# Multiplexed Detection of Sepsis Markers in Whole Blood using Nanocomposite Coated Electrochemical Sensors

**DOI:** 10.1101/2020.11.03.20224683

**Authors:** Uroš Zupančič, Pawan Jolly, Pedro Estrela, Despina Moschou, Donald E. Ingber

## Abstract

Sepsis is a leading cause of mortality worldwide that is difficult to diagnose and manage because this requires simultaneous analysis of multiple biomarkers. Electrochemical detection methods could potentially provide a way to accurately quantify multiple sepsis biomarkers in a multiplexed manner as they have very low limits of detection and require minimal sensor instrumentation; however, affinity-based electrochemical sensors are usually hampered by biological fouling. Here we describe development of an electrochemical detection platform that enables detection of multiple sepsis biomarkers simultaneously by incorporating a recently developed nanocomposite coating composed of crosslinked bovine serum albumin containing a network of reduced graphene oxide nanoparticles that prevents biofouling. Using nanocomposite coated planar gold electrodes, we constructed a procalcitonin sensor and demonstrated sensitive PCT detection in undiluted serum and clinical samples, as well as excellent correlation with a conventional ELISA (adjusted r^2^ = 0.95). Sensors for two additional sepsis biomarkers — C-reactive protein and pathogen-associated molecular patterns — were developed on the same multiplexed platform and tested in whole blood. Due to the excellent antifouling properties of the nanocomposite coating, all three sensors exhibited specific responses within the clinically significant range without any cross-reactivity in the same channel with low sample volume. This platform enables sensitive simultaneous electrochemical detection of multiple analytes in human whole blood, which can be expanded further to any target analyte with an appropriate antibody pair or capturing probe, and thus, may offer a potentially valuable tool for development of clinical point-of-care diagnostics.

**GRAPHICAL ABSTRACT:** 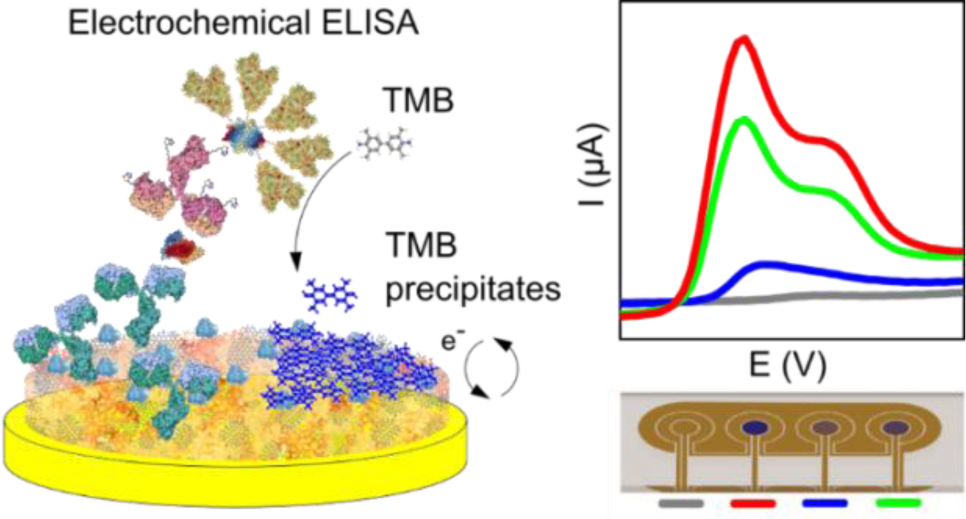

---

Specific and sensitive quantification of multiple biomarkers for point-of-care (POC) diagnostics applications using whole blood samples has been a holy grail in clinical diagnostics for decades. The greatest need for such devices is the diagnosis of life-threatening medical conditions, such as sepsis, where every hour of delay in administration of the correct antibiotic increases the mortality rate by over 7%.^1^ Unfortunately, no single sepsis biomarker exhibits appropriate specificity and sensitivity for disease diagnosis, and thus multiplexed detection of biomarker panels has emerged as the preferred POC diagnostic option.^2^ Among the variety of biosensing platforms, electrochemical (EC) biosensors would be preferred to use for this application due to their potential for optimal detection limits, cost-effective integration with diagnostic readers, and low power instrumentation requirements.^3, 4^ However, while individual biomarker detection has been demonstrated in whole blood using electrochemical systems,^5, 6^ it only has been possible to carry out multiplexed detection by prefiltering the blood, capturing and then releasing the biomarker molecules,^7^ which adds greatly to the complexity of a potential integrated ‘sample in-answer out’ device. Commercial EC POC platforms have been developed that utilize separate cartridges for different biomarkers (e.g., Abbott’s i-STAT system), but they do not carry out multiplexed protein detection and quantification cartridge.^8^ Therefore, there is a need for a low-cost and straightforward multiplexed EC platform that can rapidly quantify multiple biomarkers in a complex sample, such as serum, plasma, or whole blood.

One of the reasons behind the limited commercial success of affinity-based EC sensors in complex biological samples is the high susceptibility of the EC sensing elements to biological fouling.^9^ Nonspecific binding can drastically impact resistive and capacitive properties of the sensing element, hampering electrochemical signal transduction.^10^ We recently demonstrated that this issue can be addressed by coating electrodes with a three-dimensional (3D) nanocomposite containing crosslinked bovine serum albumin (BSA) doped with conducting nanomaterials, such as gold nanowires (AuNWs) or carbon nanotubes (CNTs), and using 3,3′,5,5′-tetramethylbenzidine (TMB) that precipitates locally on the surface of the electrode to quantify binding.^11, 12^ This matrix allowed excellent electrochemical signal transduction between the gold electrode and the nanocomposite surface, which could be functionalized with antibodies while maintaining conductive and antifouling surface properties.^11, 12^ Although the nanocomposite demonstrated excellent antifouling characteristics by maintaining its properties after one-month exposure to human plasma, the addition of expensive AuNWs to the matrix can pose a significant challenge towards commercialization of this technology. Thus, in the present study, we replaced AuNWs with reduced graphene oxide nanoflakes (rGOx) with known antifouling and electrochemical attributes, which are also much less expensive.^13, 14^ By doing so, the cost of the nanocomposite was reduced by ∼99%, which should enable this technology to have an immediate commercial impact. The EC sensor platform we present here, which is enabled by the graphene oxide nanoflakes-based antifouling nanocomposite, can quantify multiple biomarkers and can be easily incorporated with microfluidics. An antibody-based sensor for one of the most important sepsis biomarkers, procalcitonin (PCT), is characterized in serum and whole blood, and the sensor is validated using clinical samples. The broad applicability of the nanocomposite is further demonstrated by the incorporation of two additional sensors for two other infection biomarkers that are not based on antibodies as capture probes: C-reactive protein (CRP),^15^ which is based on use of a phosphorylcholine (PC) capturing probe, and pathogen-associated molecular patterns (PAMPs) that are bound by immobilized Fc-Mannose Binding Lectin (Fc-MBL).^16^ This is the first time a multiplexed EC sensor for these three different sepsis biomarkers has been demonstrated in whole blood.

## RESULTS AND DISCUSSION

### Nanocomposite characterization

Nanocomposite coatings composed of BSA and AuNWs cross-linked with glutaraldehyde (GA) have been shown to demonstrate excellent antifouling properties, maintaining high conductivity even after one-month exposure to human plasma and allowing sensitive detection of interleukin-6 (IL-6).^17^ To lower the cost of the nanocomposite, AuNW were replaced with conducting rGOx nanoflakes that were terminated with either a carboxyl or amine group. We first constructed an EC sensor using this modified nanocomposite coating and analyzed its ability to detect IL-6 in undiluted serum to compare its sensitivity relative to the AuNW sensor studied in the past and confirm its functionality. The EC sensor employing the nanocomposite with carboxyl terminated rGOx and use of precipitating TMB resulted in a limit of detection (LOD) of 63.1 pgmL^-1^ for IL-6 in undiluted human serum, while the amine-terminated rGOx was even more sensitive (LOD = 13.6 pgmL^-1^) (**Figure S1**). Importantly, the latter result represents a significant improvement relative to our previous described IL-6 sensor using the AuNW containing composite (LOD = 23 pgmL^-1^).^11^ A high concentration of labeling molecules contributes to high sensitivity, and we found that concentrations of streptavidin-polymerized HRP conjugate (Strep-PolyHRP) as high as 1 µgmL^-1^ could be used successfully for target labeling with minimal non-specific binding (**Figure S2**).

Covalent interaction between the conducting nanomaterial and the BSA matrix improves the efficiency of nanomaterial incorporation, allows for high dispensability, and allows the composite to maintain its structure over time,^18^ maintaining its antifouling properties. Altering the terminal groups of rGOx affects its solubility and agglomeration in solution, and hence the distribution of the nanoflakes within the nanocomposite and its final stability.^19^ Amine terminated rGOx nanoflakes allow covalent linking of the nanomaterial within the nanocomposite through glutaraldehyde pyridine polymers that are also responsible for crosslinking of the BSA,^11^ which may contribute to the increased sensitivity we observed with these coatings.

Field emission electron scanning microscope (FE-SEM) imaging of the GA crosslinked BSA/rGOx coating deposited on the gold surface revealed that it is a densely packed composite (**Figure 1A**). At the same time, use of atomic force microscopy (AFM) to analyze the film’s porosity revealed higher surface roughness (RMS = 1.290 nm) when compared to gold with no nanocomposite that underwent the same treatment (RMS = 731.1 pm) (**Figure 1B** and **Figure S3**) confirming the formation of the cross-linked protein-based nanocomposite on the electrode surface. AFM topography also revealed profiles similar to previously published gold nanowire BSA composites (**Figure 1C** and **Figure S4**).

**Figure 1.**
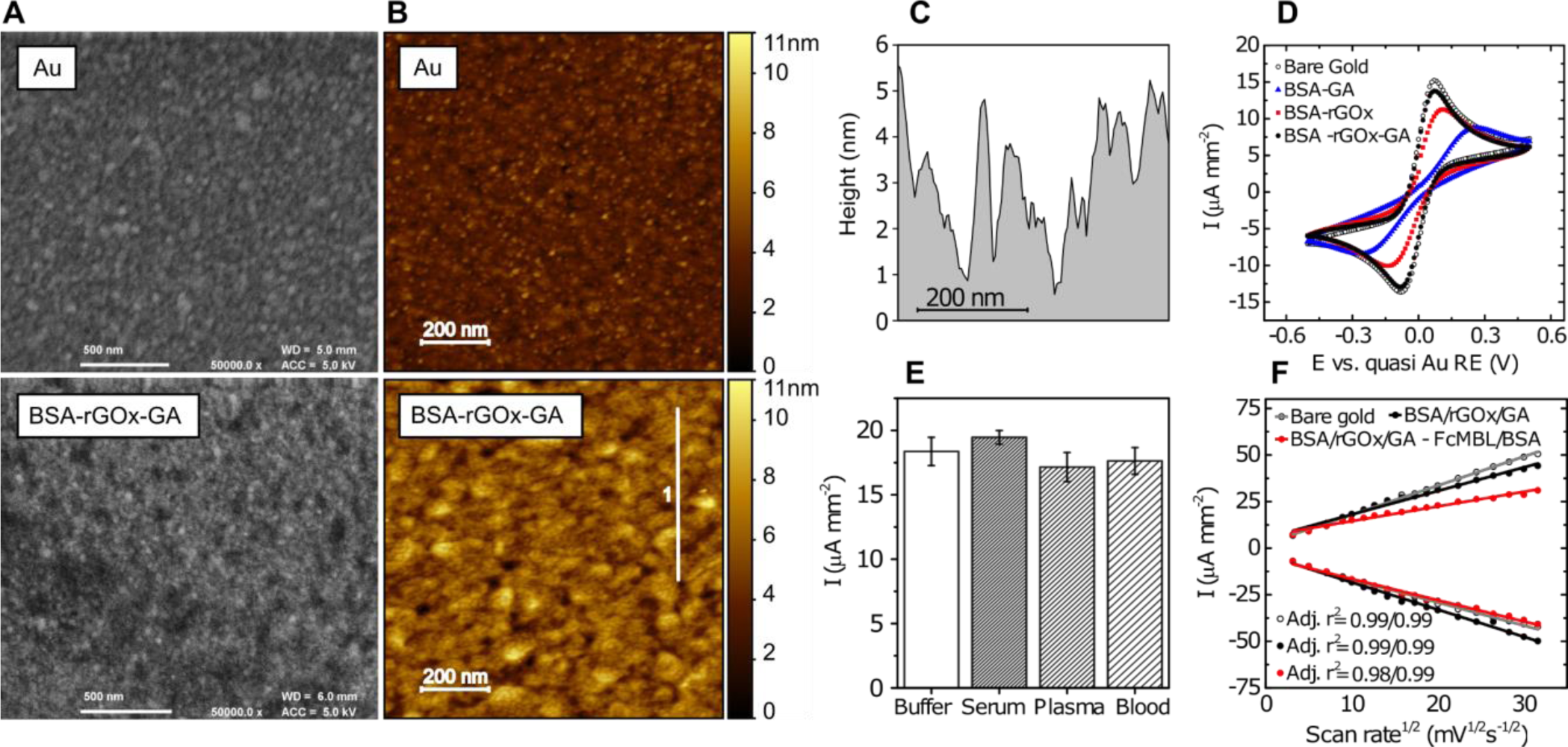
Characterization of the nanocomposite coating. (A) FE-SEM image of bare gold (Au) and nanocomposite coated gold surface (BSA-rGOx-GA). (B) AFM topography representation of bare gold (Au) and nanocomposite coated gold surface (BSA-rGOx-GA). (C) The extracted profile from the white line numbered 1 in B. (D) Cyclic voltammograms representing oxidation and reduction of 5mM ferri-/ferrocyanide solution using gold electrodes with various coatings; BSA crosslinked with glutaraldehyde (BSA-GA), BSA incorporated with rGOx nanoflakes (BSA-rGOx) and glutaraldehyde crosslinked BSA incorporated with rGOx nanoflakes (BSA-rGOx-GA). (E) Oxidation currents of ferri-/ferrocyanide couple obtained after exposure of nanocomposite coated electrodes to buffer (phosphate buffered saline, PBS), human serum, plasma, and whole blood. Bars represent the average, error bars are standard deviation of the mean, n = 4. (F) Oxidation and reduction current density of the ferri-/ferrocyanide couple against the square root of the scan rate, obtained at different stages in sensor preparation. Bare gold, nanocomposite covered gold (BSA/rGOx/GA) and Fc-Mannose Binding Lectin functionalized nanocomposite surface (BSA/rGOx/GA – Fc-MBL/BSA).

To characterize the electrical properties of the nanocomposite, cyclic voltammetry (CV) in ferri-/ferrocyanide solution was performed **(Figure 1D**). As demonstrated previously,^11^ GA cross-linked BSA results in a porous coating allowing for migration of electroactive species to the proximity of the electrode. When comparing the obtained current density of cross-linked BSA composite to a bare gold electrode, we observed 57.3% of the oxidation and 61.6% of the reduction currents, which increased when we incorporated amine-terminated reduced graphene oxide (rGOx-NH2) in the nanocomposite (respectively resulting in 74.0% and 73.7% of oxidation and reduction currents compared to bare gold). This result indicated that simply imbedding conductive rGOx flakes increase the conductivity of the surface coating; however, when BSA-rGOx-NH2 composite was covalently cross-linked with GA, the coating formed displayed 90.5% and 94.7% of oxidation and reduction currents compared to bare gold, respectively. This BSA/rGOx/GA nanocomposite maintained the same peak-to-peak separation potential (ΔEp) of 0.151 V, which was significantly higher for the BSA/rGOx composite (0.252 V) and for BSA/GA alone without embedded conducting materials (0.524 V)(**Figure 1D**).

The antifouling properties of the BSA/rGOx/GA nanocomposite were confirmed when we exposed the coated electrodes to serum, plasma, and whole blood, which are the most frequently used biological fluids used in POC testing. No significant changes in current density was observed after 60 min incubation with these complex fluids indicating minimal fouling occurred on the electrodes (**Figure 1E**). An additional benefit of the BSA/rGOx/GA nanocomposite is its versatile functionality due to the plethora of different amino acids naturally present in the BSA protein that can be used for chemical conjugation. The abundance of aspartic and glutamic acid can be explored by activation of carboxyl acid groups through carbodiimide chemistry for covalent linkage of target capture probes to the surface.

Large probes can affect surface chemistry and decrease the sensitivity of an electrochemical sensor due to their size and charge screening effect.^20, 21^ To gain an insight into the surface chemistry of the BSA/rGOx/GA nanocomposite, the genetically engineered human opsonin protein, Fc-MBL, was conjugated to the nanocomposite. The remaining activated carboxyl groups were quenched in tris buffered saline (TBS) and blocked with 0.1% BSA in TBS (**Figure 2A**). Increasing the CV scan rate in ferri-/ferrocyanide solution while analyzing the Fc-MBL-modified nanocomposite coated electrode revealed a linear relationship between the current density and the square root of the scan rate, indicating diffusion limiting oxidation and reduction of electroactive species (**Figure 1F** and **Figure S5**). Furthermore, the functionalized nanocomposite showed distinct, but small deviation from the behavior of bare gold or non-functionalized nanocomposite, indicating surface characteristics can be obtained even after the conjugation of large proteins, such as Fc-MBL (MW = 90 kD) to the nanocomposite. Previous studies using alkanethiol^22^ and polyethylene-glycol^23^ based self-assembly monolayers report considerably higher loss of electroactivity after surface functionalization with capturing probes.

**Figure 2.**
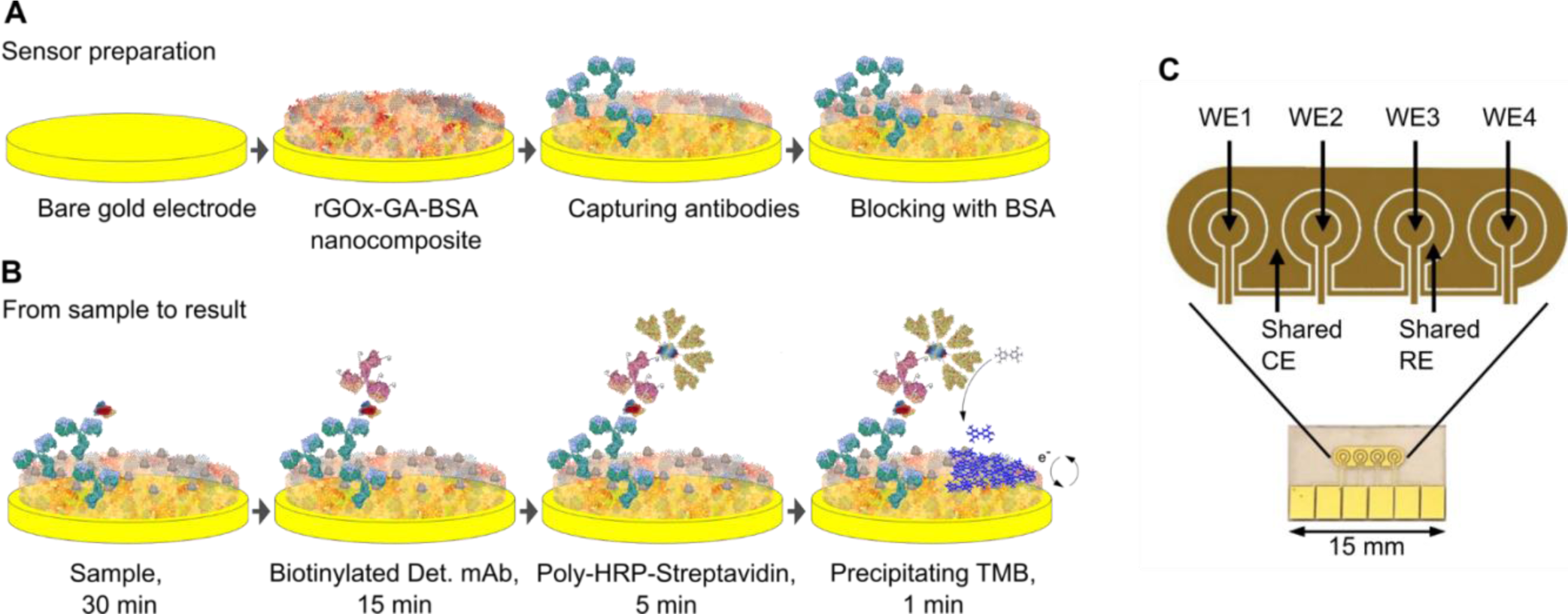
Electrochemical ELISA. (A) Preparation of the electrochemical sensor. (B) A stepwise process involving the application of the sample, biotinylated detection antibody, polymerized conjugate of streptavidin and horseradish peroxidase, and precipitating TMB. Between each step, an additional washing process is introduced (see Methods). (C) Planar evaporated gold electrode chip comprising of four individual working electrodes (WE 1 - 4), shared counter electrode (CE) and shared quasi reference electrode (RE) allowing for four separate sensors to be constructed in an area smaller than 7 x 2.5 mm.

### The multiplexed electrochemical sensor platform

We then leveraged the excellent electrochemical properties of the nanocomposite to construct a sensitive electrochemical ELISA using the coated electrode (**Figure 2B**). Due to the ultra-low fouling, non-specific interactions are minimized when samples come in contact with the surface. The biotinylated detection mAb and a conjugate of Strep-PolyHRP were introduced in separate steps to allow for greater versatility during assay development. In the final step precipitating TMB was used, which precipitates locally to the electrode where the enzyme is present. The sensor output was read by cyclic voltammetry at 1 V/s to ensure complete oxidation of the precipitated TMB on the surface. Local precipitation of the TMB enables the construction of a multiplexed sensing array where multiple electrodes with various capturing probes can be placed in close proximity. To demonstrate proof-of-principle, we constructed a sensor with 4 gold working electrodes that shared a pseudo reference gold electrode and counter electrode, all compacted in an area less than 7 x 2.5 mm (**Figure 2C**).

### Procalcitonin detection in complex biological fluids

Procalcitonin, an FDA approved biomarker used for assessing risk of developing sepsis in critically ill patients^24^ rises from 50 pg mL^-1^ to over 10 ng mL^-1^ in cases of fulminant sepsis.^25, 26^ An electrochemical sensor for PCT was constructed on top of the nanocomposite using anti-PCT monoclonal antibody (mAb), as described above. Step by step validation of the assay construction was confirmed by surface plasmon resonance (SPR) analysis, which confirmed specific binding of PCT to the capturing mAb and its binding to biotinylated secondary mAb, as well as Strep-PolyHRP binding to biotin moieties (**Figure S6**). When the sensor was tested in buffer (**Figure S7**), undiluted serum, and 50% diluted whole blood (**Figure 3A**) using various PCT concentrations spanning four orders of magnitude, we observed similar behavior in all three fluids. In undiluted serum, the PCT sensor revealed a large dynamic range (0.09 – 10.24 ng mL^-1^) and an LOD of 64.5 pg mL^-1^. When tested in 50% diluted blood, the sensitivity of the sensor surprisingly increased (LOD of 24.7 pgmL^-1^), but this was accompanied by a decrease in a dynamic range (0.07 – 2.49 ngmL^-1^). The dissociation constant (Kd = 0.45 ± 0.07 ngmL^-1^) measured in diluted blood also remained comparable to the sensor in buffer (Kd = 0.42 ngmL^-1^ ± 0.07 ngmL^-1^). With increasing PCT concentration, the area of TMB oxidation peaks increased (**Figure 3B**), and two TMB oxidation peaks could be distinguished due to the excess TMB precipitated on the surface when abundant amounts of enzyme are present. A thick layer of TMB can have an insulating effect,^10^ and higher potentials are needed to complete the oxidation of TMB that is present further away from the surface.

**Figure 3.**
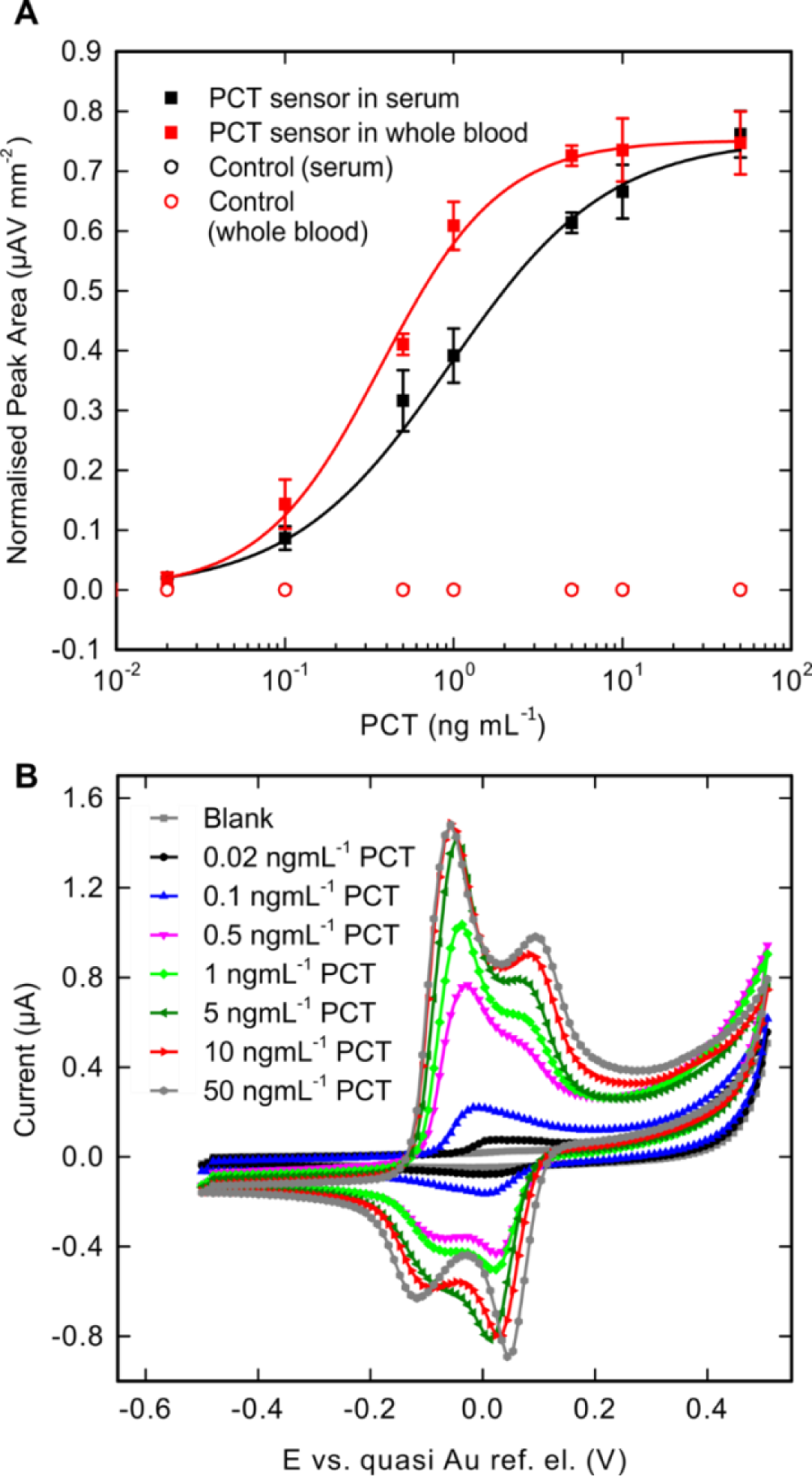
(A) PCT sensor response in undiluted serum (black) and 50% diluted whole blood (red). Squares represent the average (n = 3) and error bars the standard deviation. Hill fit was performed to fit the data. Empty circles represent the response obtained using the control electrode with no capturing mAb. (B) TMB oxidation peaks obtained with a CV from PCT sensor in whole blood with a blank and increasing PCT concentrations.

In every experiment, one electrode was not modified with PCT mAb, and this was used as an on-chip negative control. Analysis of the neighboring electrodes revealed that no current peaks were observed in the electrode without capturing mAb even at highest PCT concentration (**Figure S8**), again demonstrating the potent antifouling properties of the nanocomposite surface coating and the highly localized precipitation of catalyzed TMB. The main advantage of POC devices in the field of infectious diseases is the time it takes from the start of the analysis until the result.^27^ Importantly, the turnaround time of this prototype EC sensor was 51 min, which meets the clinical need of diagnosis within the first hour.^1^

### Microfluidic integration of the PCT sensor

To further reduce the analysis time, the EC sensor chip was coupled with a poly-dimethylsiloxane (PDMS) block containing a microfluidic channel using double sided adhesive tape, and detection of PCT in serum was performed under flow at 10 µL/min with a sample size of 50 µL (**Figure 4E**rror! Reference source not found.**A**). The implementation of this microfluidic sensor assay reduced the total time of sample and label incubation to 7 min. Again, the antifouling coating performed well under flow with minimal cross reactivity demonstrated by the on-chip electrode control (**Figure 4**Error! Reference source not found.**B**). These findings demonstrated the coating can be used with microfluidic systems and that it retains its properties under conditions involving increased diffusion. The data obtained with this PCT electrochemical sensor were fitted using a Hill fit equation, which resulted in determination of a Kd of 8.69 ± 3.85 ngmL^-1^ with a dynamic range of 1.06 – 48.8 ngmL^-1^.

**Figure 4.**
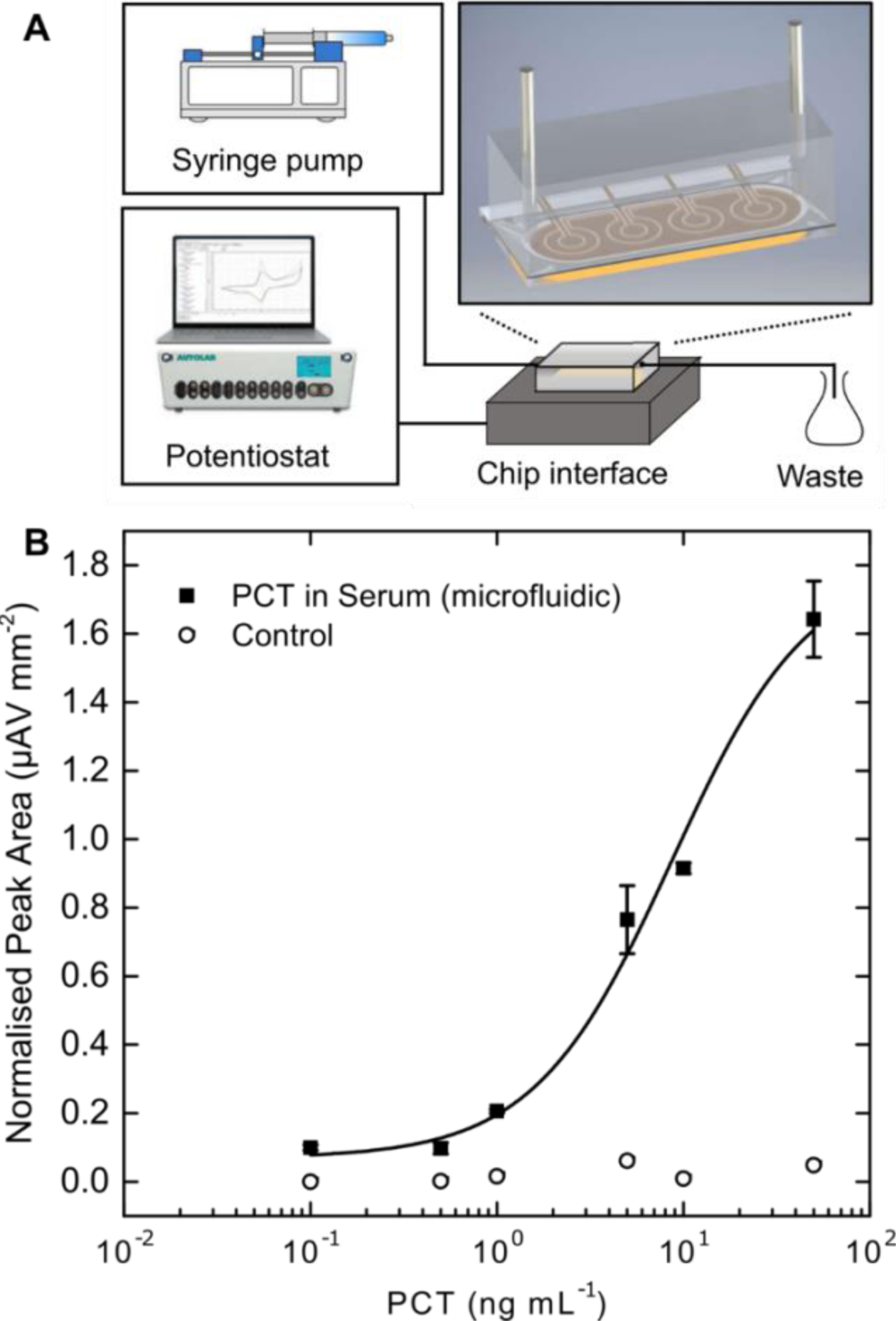
Microfluidic integration of the PCT sensor. (A) Schematic representation of microfluidic integration by incorporation of nanocomposite covered planar electrode to a PDMS with the channel outlined by double sided adhesive, connected to a syringe pump and a potentiostat through a chip interface. (B) Response curve obtained with 50 µL sample using flow of 10 µL/min decreasing the analysis time below 10 min. Squares represent the average (n = 3) and error bars the standard deviation. Empty circles represent the response obtained using the control electrode with no capturing mAb.

### Clinical study

To further validate the PCT sensor we analyzed 21 patient serum samples obtained as part of a past clinical study on detection of blood stream infections^16^ using the non-microfluidic integrated protocol described above (**Figure 3**). The data obtained were compared to the results acquired using a conventional plate ELISA (1:10 sample dilution was used due to the smaller linear range of the plate-based ELISA). As the electrochemical sensor analysis was performed using undiluted serum where PCT levels can increase up to 100 ng mL^-1^,^28^ responses in 9 samples were out of range of the EC sensor. However, the remaining 12 samples were quantified using both methods, and concentrations obtained with plate ELISA and electrochemical sensors were compared. The values obtained with both methods were plotted in a scatter plot (**Figure 5A**) and linear regression analysis revealed an adjusted r^2^ of ∼0.95 representing excellent correlation between the electrochemical PCT sensor and conventional ELISA. There also was excellent agreement between the two methods in the lower concentration region (below 1 ngmL^-1^) when a Bland-Altman plot was used (**Figure 5B**); this is especially important when the EC sensor is used for early sepsis diagnosis, as higher PCT levels are corelated to more severe sepsis.^29^ It is worth mentioning that higher variability at higher concentrations was observed which is likely due to saturation of EC sensor.

**Figure 5.**
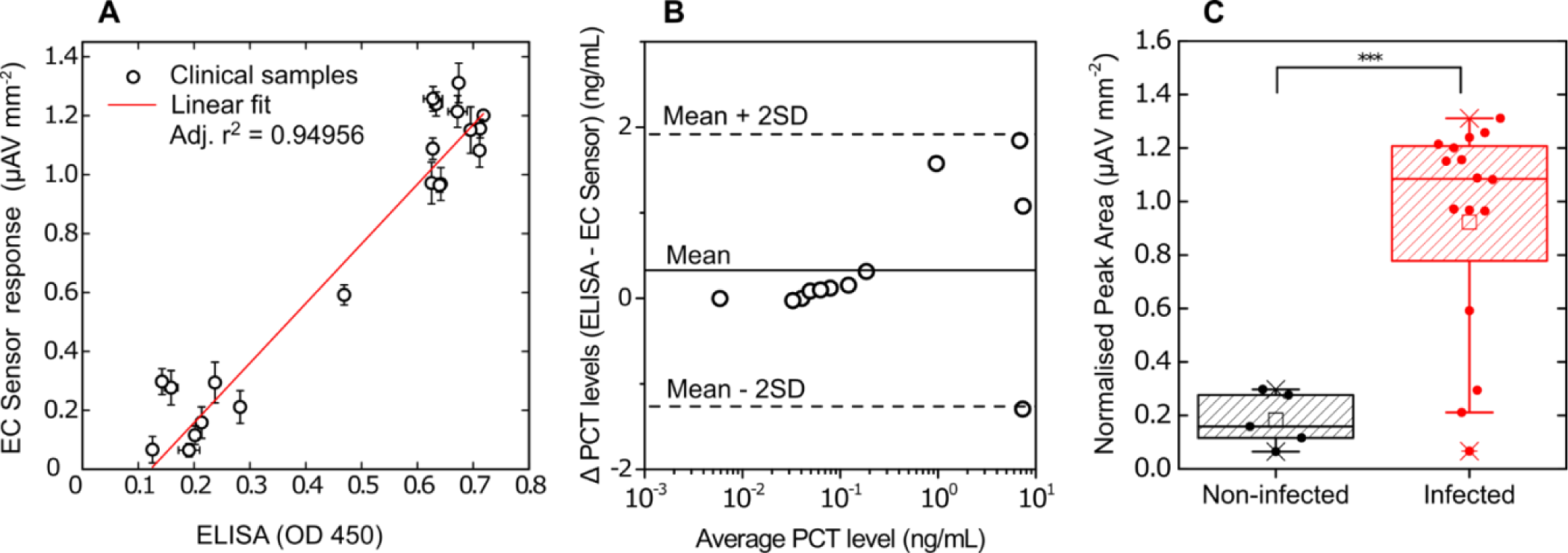
Evaluation of electrochemical PCT sensor in a clinical study. (A) Electrochemical (EC) sensor response vs. ELISA response using sepsis patients’ and non-infected patients’ clinical samples. Circles represent the mean and error bars represent the standard deviation, n = 3 (EC sensor) and n = 2 (ELISA). Red line represents the linear fit. (B) Bland-Altman plot describing the agreement between EC sensor and plate ELISA. (C) Box plot comparing PCT levels in infected and non-infected patient groups obtained with EC sensor. Full circles represent the mean of the measured values, lines in a box represent the median, 25^th^ and 75^th^ percentile, empty square is the mean value, whiskers represent 10^th^ and 90^th^ percentile and *\*\*\** represents p < 0.001.

When grouping the clinical samples into infected and non-infected patient groups, both the electrochemical sensor and conventional ELISA successfully distinguished between the two groups with high confidence; however, some positive sepsis samples still exhibited low PCT levels in both assays (**Figure 5C** and **Figure S9**), clearly demonstrating the disadvantage of using a single biomarker for sepsis diagnosis. This is a common problem with the PCT assay in clinical settings, ^30, 31^, including in the past study using the same clinical samples with another PCT assay^16^; the PCT assay is also not highly specific.^32^ Thus, current recommendations do not recommend the use of PCT as a single biomarker for prognosis, and clearly state the need to use multiple clinical biomarkers for sepsis diagnosis.^33-35^

### Multiplexed detection of PCT, CRP and PAMPs for sepsis diagnosis

The use of multiple biomarkers for sepsis diagnosis can increase the test accuracy, however the biomarkers must be chosen carefully.^36, 37^ The highest value of the biomarker panel can be achieved by combining assays for indirect infection protein biomarkers, such as PCT and CRP with direct infection markers that measure the presence of pathogens in the blood sample.^38^ This latter goal of direct detection of pathogens in blood is no trivial task, but we have previously shown that it can be achieved using the Fc-MBL opsonin protein that binds to over 100 different pathogens of all classes (gram −/+ bacteria, viruses, fungi, parasites) as well as pathogen-associated molecular patterns^39, 40^, such as lipopolysaccharide endotoxin and lipoteichoic acid, which are present on the surfaces of pathogens and released into blood when the pathogens are killed.^39, 40^ Importantly, using a Fc-MBL PAMPs capture assay, we have previously shown that the presence of pathogens can be detected within patient whole blood within 1 hour after collection with high sensitivity (>81%), specificity (>89%), and diagnostic accuracy (0.87), even in patients that are blood culture negative.^16^

We also used human whole blood in the current study because no processing steps are needed after sample collection, which eliminates the risk of removing biomarkers and potential pathogens from patient samples. The previously described protocol for PAMPs capture by Fc-MBL^40^ was adapted and the sensor was tested in buffer (**Figure S10**) as well as 50% diluted whole blood (**Figure 6A**) using mannan as a standard. The electrochemical sensor in whole blood exhibited higher currents at low mannan levels than buffer due to the baseline presence of low levels of PAMPs in blood, although the dissociation constants were similar (9.3 ± 0.9 versus 5.1 ± 3.0 ng mL^-1^ for buffer and blood, respectively). Similar observations were made by comparing buffer and blood-based analysis using an Fc-MBL plate-based ELISA (**Figure S11**), which again demonstrated successful transfer of the Fc-MBL-based assay to an electrochemical platform. Standard deviations were higher in blood as expected, which led to lower sensitivity when compared to sensing in buffer (LOD = 6.0 versus 4.1 ng mL^-1^); however, the dynamic range in blood was improved, which is important for biomarker quantification.

**Figure 6.**
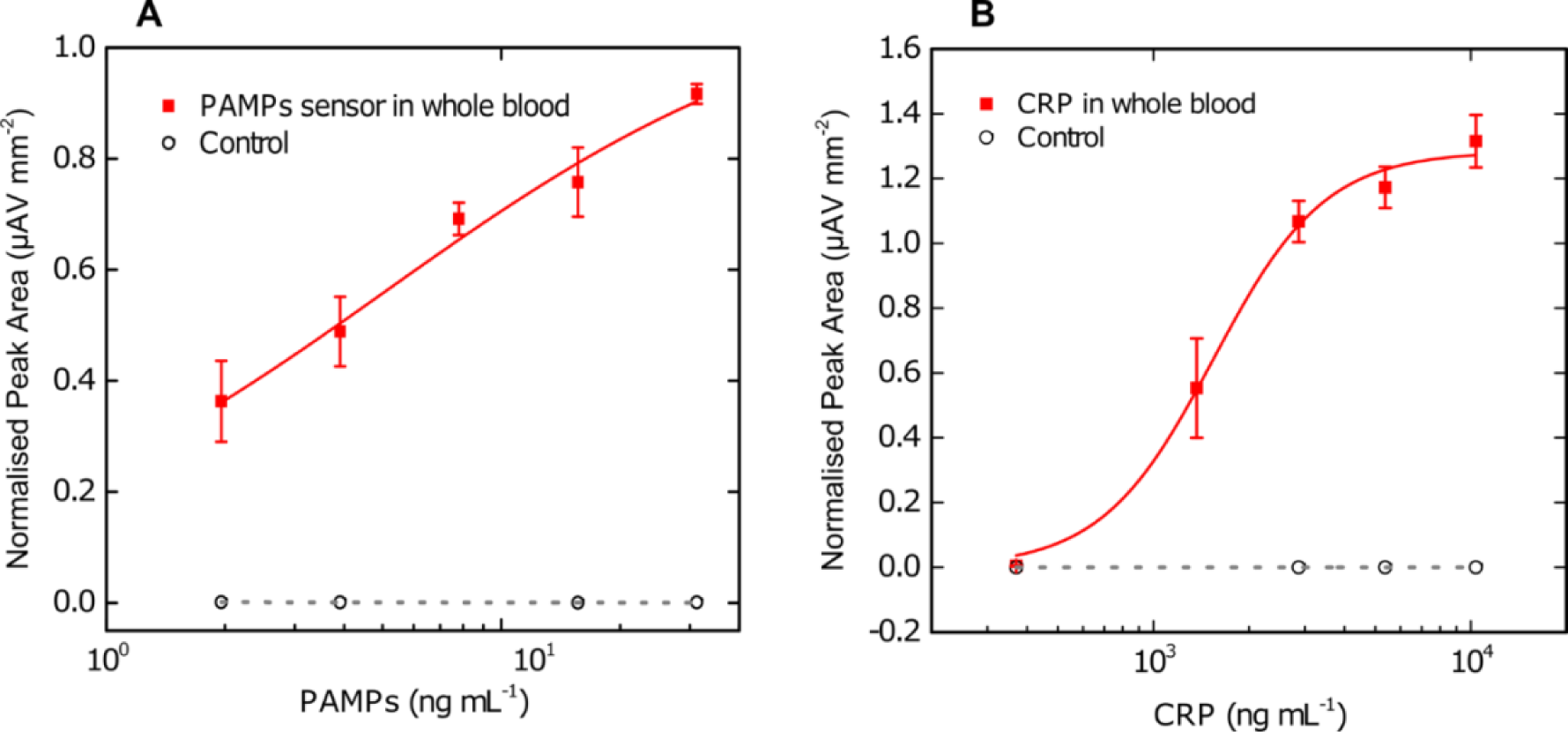
PAMPs sensor based on Fc-MBL technology and CRP sensor based on phosphocholine. (A) PAMPs sensor response in 50% diluted whole blood. Mannan is added to the matrix to obtain the calibration curve. (B) CRP sensor in 50% diluted whole blood. Squares represent the average (n = 3) and error bars the standard deviation while empty circles represent the response obtained using the control electrode with no capturing probe in A and B. Hill fit was performed to fit the data in both experiments.

To further increase the sensitivity and accuracy of the potential electrochemical sepsis diagnostic sensor, a CRP assay was constructed and integrated into the same platform with the PCT and Fc-MBL-based PAMPs sensors. Plasma CRP levels can increase to over 100 µgmL^-1^ in septic patients and the recommended cut-off values are highly variable. For example, a cut-off value for diagnosis of infection adults has been proposed to be 79 µg mL^-1^,^41^ while for neonatal sepsis it is 4.82 µg mL^-1^.^42^ Detection of CRP in parallel to PCT is also challenging, as concentrations of CRP are three to five orders of magnitude higher than PCT. Thus, to perform measurements of the CRP in whole blood in the physiological range and obtain a calibration curve, we depleted the CRP content in whole blood from 1.44 µgmL^-1^ to 0.37 µgmL^-1^ with the use of biotinylated anti-CRP mAbs bound to streptavidin coated magnetic beads, which we used to capture the CRP protein and thereby reduce its levels in blood samples (**Figure S12**).

We first tacked this challenge relating to large differences in concentrations of these two biomarkers in blood by developing a mAb based sensor in which the secondary antibody was conjugated directly to a single HRP moiety, hence avoiding large signal amplification through the Strep-polyHRP conjugate that we used for PCT and Fc-MBL (**Figure S13**). Although smaller currents were observed, the sensor was already saturated at 0.37 µgmL^-1^ of CRP, hence an alternative approach using a low affinity capturing probe was used. Phosphocholine (PC) is known to bind to CRP ^43, 44^ in the presence of calcium ions and its dissociation constant is 5 μM at a physiological Ca^2+^ concentration (10 mM);^45^ this is in contrast to most antibodies that have dissociation constants within the nM range.^46^ In our sensors, we conjugated a PC derivative, 4-aminophenyl-phosphorylcholine (APPC) to the nanocomposite through EDC/NHS chemistry.

When this electrochemical sensor was used to detect CRP in buffer (**Figure S14**), we still observed a response in the nM range indicating strong signal amplification through the Strep-PolyHRP part of the assay with LOD at 10 ngmL^-1^ and a Kd of 55.7 ± 4.4 ngmL^-1^. Performing EC CRP assay in CRP-depleted blood diluted 1:1 with buffer allowed us to obtain a calibration curve that was significantly shifted when compared to the assay in buffer alone (**Figure 6B**). This may be the consequence of CRP binding to phosphocholine groups in blood lipoproteins and phospholipids in the membrane of blood cells,^44^ which would compete with the PC probes on the electrode surface for the CRP binding pocket. Our studies revealed that the LOD of the final CRP electrochemical assay was 0.492 µgmL^-1^, with a Kd = 1.54 ± 0.1 µgmL^-1^ and a dynamic range of 0.63 – 3.76 µg mL^-1^; this shifted the sensor range from ng mL^-1^ to µg mL^-1^, which enabled direct measurement of CRP in blood.

To demonstrate the three sensors can be multiplexed within a single chip, electrodes were individually functionalized with each capturing probe. Each analyte was added at the highest concentration according to its respective calibration curve, and exposed to the sensor in buffer (**Figure 7**). Minimal cross-reactivity was observed in three independent experiments for each condition demonstrating the nanocomposite technology can be used to create multiplexed electrochemical sensors with multiple antibody pairs, as long as there is no cross-reactivity between the antibodies.

**Figure 7.**
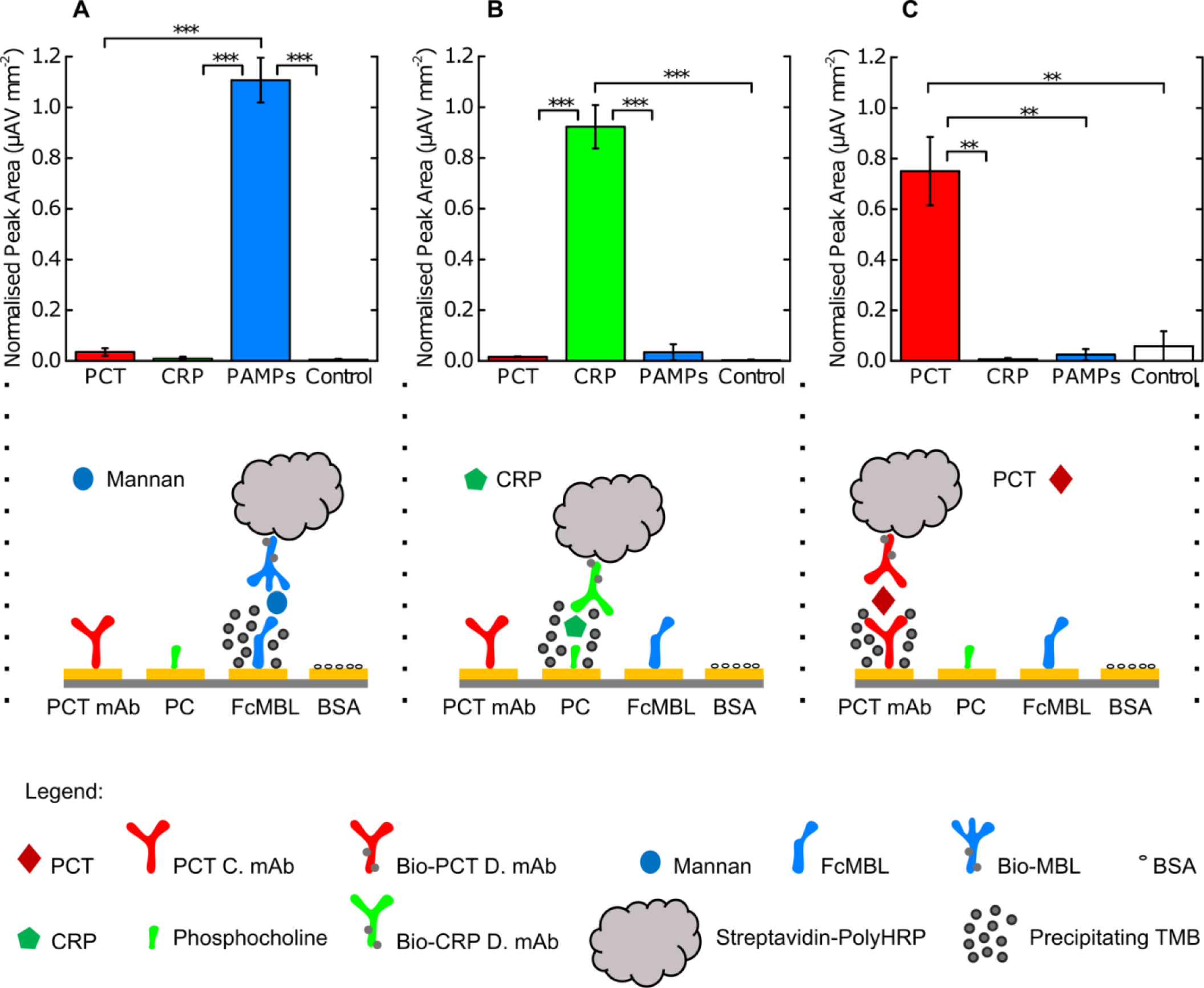
Cross-reactivity of PAMPs, CRP and PCT sensors on the same chip. Chips comprising of PCT, PAMPs, CRP and control sensors on WE1, WE2, WE3 and WE4 were exposed to 31.25 ngmL^-1^of mannan (A), 10 µg/mL^-1^ of and CRP (B) and 50 ngmL^-1^ of PCT (C). Bars represent the mean value and error bars the standard deviation (n = 3). *\*\*\** represents p<0.001 while *\*\** represents p<0.01 in a two-sample T-test. The schematic below represents the probe on the sensors surface (PCT mAb, PC, Fc-MBL and BSA for control) with the addition of respected targets, leading to specific response of the EC sensor. C. mAb refers to capturing mAb, bio-PCT/CRP D. mAb/MBL refers to biotinylated detection PCT/CRP mAb and biotinylated MBL.

To confirm parallel detection of the three analytes using these multiplexed electrochemical sensors, four chips were constructed and tested as follows. The first chip was exposed to 50% diluted blood (with depleted CRP levels) with no additional analytes; the second was exposed to mannan; the third to mannan and CRP; and the final chip to mannan, CRP and PCT, all in whole blood diluted 1:1 with buffer. An increase in current density was observed where the analytes were added, and the overall signature of each electrochemical sensor changed based on the addition of its respective analyte (**Figure 8**). We observed a response in the PAMPs sensor only with blood, which could be associated with presence of PAMPS in the blood constituting the baseline PAMPs level (**Figure 6a**). Addition of mannan, which is the ligand for Fc-MBL found in PAMPS, only increased the signal of the Fc-MBL based sensor. When CRP and mannan were added, both the CRP and PAMP sensors responded, and upon addition of all analytes, all sensors responded accordingly.

**Figure 8.**
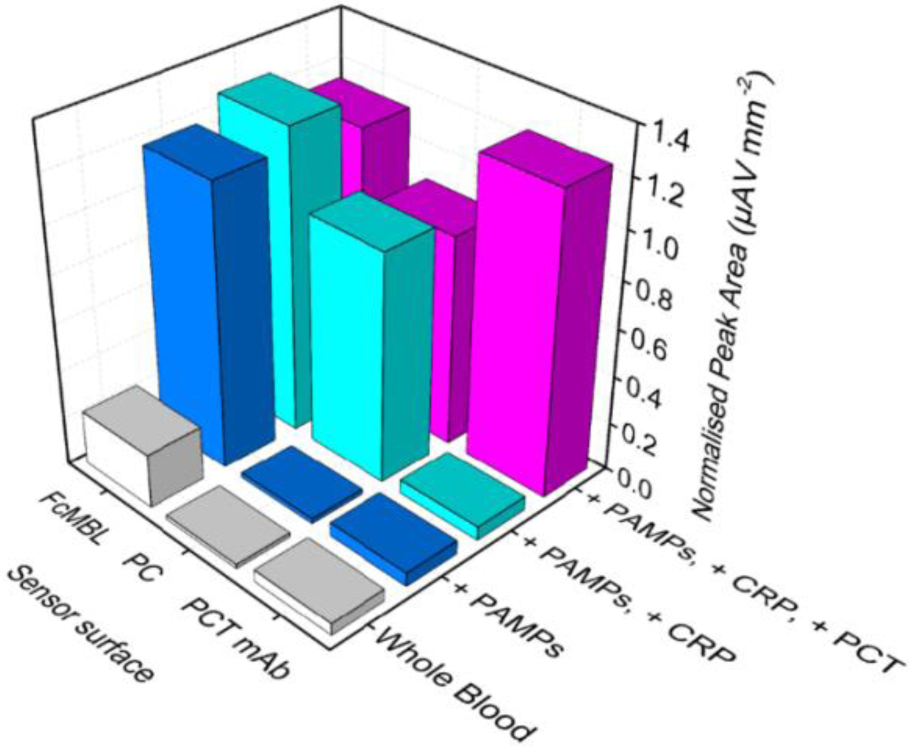
Multiplexed detection of PCT, CRP and PAMPs in whole blood. Chips comprised of three sensors were exposed to blood with lowered CRP levels, blood with spiked mannan (31.25 ngmL^-1^), blood sample with mannan and CRP (10 µgmL^-1^) and a blood sample with mannan, CRP and PCT (50 ngmL^-1^).

## CONCLUSION

In conclusion, we demonstrated that conductive, amine-terminated rGOx nanoflakes can be successfully integrated within a previously described BSA-based nanocomposite and cross-linked with GA to form a porous conductive layer for electrodes exhibiting excellent antifouling properties. This nanocomposite significantly lowered the cost and improved the sensitivity of IL-6 sensors compared to a previous sensor coated with the same nanocomposite containing AuNWs rather than rGOx. The nanocomposite is highly versatile as it can be functionalized with analyte-capturing probes ranging from antibodies and engineered proteins to small molecules (e.g., APPC) to construct affinity-based electrochemical sensors for detection of virtually any molecular analyte. Detection of an important sepsis biomarker (PCT) was demonstrated in buffer, serum and 50% diluted whole blood, and the sensor was validated in a clinical study using patient blood specimens. The electrochemical sensor was also integrated into a microfluidic assay and detection of PCT in undiluted serum was demonstrated in less than 10 minutes indicating the potential value of this technology for POC applications. In addition, we constructed a PC-based CRP sensor that enabled simultaneous detection of PCT and CRP in the same sensor chip with no cross-reactivity, even though the first biomarkers is present at ng mL^-1^ concentrations, while the latter is in the µgmL^-1^ range. We also constructed an electrochemical Fc-MBL based PAMPs sensor and demonstrated its utility in whole blood for the first time.

Finally, we multiplexed all three sensors (PCT, CRP, and PAMPs) on the same electrochemical platform to develop a prototype sepsis diagnostic test which showed no cross-reactivity enabling parallel multiplex detection of their respective analytes in whole blood. Due to the use of a sandwich ELISA and TMB substrate that precipitates locally with high accuracy, sensors for multiple different proteins can now be constructed in close proximity, which allows for creation of multiplexed sensor arrays on miniaturized biosensing chips. When coupled with microfluidics, these novel multiplexed sensors could detect a panel of biomarkers in whole blood in a matter of minutes, opening the way to low-cost and easy-to-use rapid sepsis diagnostics. As a platform technology, these nanocomposite-enabled sensors also may revolutionize POC diagnostics for a broad range of other diseases and conditions.

## METHODS AND EXPERIMENTAL SECTION

### Chip preparation

Gold chips were prepared using standard photolithography process by depositing 20 nm of titanium and 150 nm of gold on a glass wafer, as described previously.^11^ Prior to use, gold chips were cleaned by 5 min sonication in acetone (Sigma Aldrich, USA, no. 650501) and then in isopropanol (Sigma Aldrich, USA, no. W292907). To ensure a clean surface, the chips were then treated with oxygen plasma using a Zepto Diener plasma cleaner (Diener Electronics, Germany) at 0.5 mbar and 50% power for 2 min.

### Nanocomposite preparation

Amine-functional reduced graphene oxide (Sigma Aldrich, USA, no. 805432) was mixed with 5 mgmL^-1^ BSA (Sigma Aldrich, USA, no. 05470) in PBS solution (Sigma Aldrich, USA, no. D8537), ultrasonicated for 1 h using 1 s on/off cycles (Bransonic, CPX 3800), heated at 105°C for 5 min to denature the protein and centrifuged to remove the excess aggregates. The nanomaterial solution was then mixed with 70% glutaraldehyde (Sigma Aldrich, USA, no. G7776) for crosslinking in the ratio of 69:1, deposited on the glass chip with gold electrodes and incubated in the humidity chamber for 20-24 h to form a conductive nanocomposite.

### Nanocomposite imaging

The nanocomposite was prepared on gold coated glass SPR chips (Reichert Technologies, Germany, no. 13206060). The images of the nanocomposite on gold were taken using a JEOL JSM-6301F field emission scanning electron microscope at 5 kV acceleration and 50000x magnification. AFM was used to evaluate the surface topography and roughness with a Digital Instruments Nanoscope IIIA AFM and Gwyddion software was used for image processing.

### Nanocomposite electrochemical characterization

Cyclic voltammograms (CV) characterizing the conductive nanocomposite were obtained in 1xTBST (Tris buffered saline with Tween 20) with 5 mM CaCl2 with 5 mM [Fe(CN)6]^3-/4-^ at a 100 mVs^-1^ scan rate using chip-integrated gold counter and gold quasi-reference electrodes. Nanocomposite antifouling properties were characterized by incubating nanocomposite coated chips in PBS buffer, human serum (Sigma Aldrich, USA), human plasma (Sigma Aldrich, USA) and whole blood (Research Blood Components, USA) for 60 min and washed for 5 min with PBS and analyzed by cyclic voltammetry in PBS with 1 M KCl and 5 mM [Fe(CN)6]^3-/4-^ at 100 mVs^-1^.To characterize the reversibility of the system, bare gold electrodes, electrodes covered with nanocomposite and electrodes with functionalized nanocomposite (*i*.*e*. with covalent attachment of PAMPs probe FcMBL and surface blocking with BSA) were analyzed by CV in 1xTBST with 5 mM CaCl2 with 5mM [Fe(CN)6]^3-/4-^ by increasing the scan rate from 10 to 1000 mVs^-1^.

### Preparation of PCT, CRP and PAMPs sensors

After nanocomposite preparation, gold chips were washed in PBS by agitation (500 rpm) for 30 min and dried with pressurized air. 1-ethyl-3-(3-dimethylaminopropyl) carbodiimide hydrochloride (Thermo Fisher Scientific, USA, no. 22980) and N-hydroxysuccinimide (Sigma Aldrich, USA, no. 130672) were dissolved in 50 mM MES buffer (pH 6.2) at 400 mM and 200 mM, respectively and deposited on nanocomposite covered gold chips for 40 min. After surface activation, chips were quickly rinsed with ultra-pure water, dried and the capturing probe was spotted on top of the working electrode area. For PCT sensor, PCT capture mAb (Abcam, UK, no. ab222276) was diluted to 0.1 mgmL^-1^ in MES buffer. One electrode was always spotted with 0.1 mgmL^-1^ BSA as a negative control and the chips were incubated overnight in a humidity chamber. After conjugation, chips were washed and quenched in TBS (50 mM Tris) for 1 h and blocked with 0.1% BSA in TBS containing 10 mM glucose. Chips were left in the blocking solution prior to use. After this point all washing steps were done with 1 mL of the washing buffer comprising of TBST with 5 mM CaCl2. Chips were incubated with the target sample for 30 min with agitation to mimic flow. After sample incubation and the wash, 2 µgmL^-1^ of biotinylated anti-PCT detection monoclonal antibody diluted in 0.2% bovine gamma globulin in TBS with 0.05% pluronic acid and 5 mM CaCl2 was added to the chip and incubated for 15 min with agitation. Biotinylated label was then washed away and Poly-HRP-Streptavidin (Thermo Fisher Scientific, USA, no. N200) was diluted to 1 µgmL^-1^ in 0.2% bovine gamma globulin in TBS-0.05% pluronic acid with 5 mM CaCl2 and incubated on the chip for 5 min. After the wash, precipitating TMB (Sigma-Aldrich, USA, no. T9455) was incubated on the chip for 1 min and washed. Final measurement was then performed in PBST using a potentiostat (Autolab PGSTAT128N, Metrohm; VSP, Bio-Logic) by a CV scan with 1 V/s scan rate between −0.5 and 0.5 V vs on-chip integrated gold quasi reference electrode. Peak area was calculated using Nova 1.11 software and corrected with geometrical surface area of the electrode.

Antibody-based CRP sensor was constructed by functionalization of the nanocomposite with 0.1 mgmL^-1^ of anti-CRP capture mAb (R&D Systems, USA, no. DY1707) MES buffer. Separately, the same mAb was directly conjugated to HRP using Lightning-Link™ HRP labeling kit (RnD Systems, USA) and used as a label at 300 ng/mL.

The PC based sensor for CRP was composed by modifying the nanocomposite with 4-amino-phenyl-phosphorylcholine (APPC) (Toronto Research Chemicals, Canada, no. A626000) by dilution to 1 mgmL^-1^ in MES buffer for bioconjugation. Biotinylated CRP mAb (R&D Systems, USA, no. DY1707) was used as a label at 1 µgmL^-1^ in both assays. For PAMPs sensor construction, FcMBL was conjugated to the nanocomposite at concentration of 0.2 mgmL^-1^. Human MBL2 (Sino Biological, USA, 10405-HNAS) was biotinylated using EZ-Link™ NHS-PEG4-Biotin (Thermo Fisher Scientific, USA) and used as a label at 200 ngmL^-1^. All three sensors were evaluated in buffer (TBST with 5 mM CaCl2 and 0.1% BSA), undiluted serum and whole blood where target was added to the whole blood, which was then diluted 1:1 with sample buffer comprising of 100 mM heparin, 20 mM glucose, and 40 mM calcium chloride in TBST before being deposited on the sensor chip.

### Detection of procalcitonin in a microfluidic set-up

Nanocomposite preparation, antibody conjugation, surface quenching and blocking was performed as described earlier. After fitting of the PDMS block and tubing (**Figure S15**) a tube was sequentially preloaded with Strep-PolyHRP labeling solution and PCT sample in serum which was mixed with labeling mAb in a 10:1 ratio moments before tube preloading. Preloaded tube was inserted to the steel microfluidic port and flown through the sensor surface for 5 min. This was followed by a preloaded 1 min wash, 1 min incubation with Strep-PolyHRP solution and another 1min wash. 10 µLmin^-1^ flow rate was used in all experiments. Preloaded tubing was removed and TMB was added manually and incubated statically for 1 min before washing and performing the measurement in PBS.

### Clinical samples

Patient blood samples were collected at the Beth Israel Deaconess Medical Center (BIDMC) Emergency Department as part of a previous clinical study^16^. The study was approved by the BIDMC Committee on Clinical Investigations, and patients or legal designees signed a written informed consent. The study included adult septic and non-infected individuals (≥ 18 years of age) where sepsis patients were defined by meeting two or more criteria for systemic inflammatory response syndrome (SIRS) and the presence of an infection. The control group was recruited from the same department where patients did not show signs of infection nor meet the two or more SIRS criteria. The collection of control samples was approved by the Harvard University Faculty of Medicine Committee on Human Studies (protocol number M20403-101) in accordance with all national or local guidelines and regulations. Clinical serum samples were stored at −20 °C until defrosted on ice and centrifuged to remove larger aggregates. 10 µL of undiluted clinical samples were deposited on the chip and the assay was performed according to the above-mentioned protocol. Standard PCT ELISA (Abcam, UK, no. ab222276) was performed on Nunc™ MaxiSorp™ ELISA plates according to manufacturer using 10 µL of the sample diluted in 90 µL of 1% BSA in PBS. Welch t-test was used when comparing infected vs. non-infected sample groups.

### CRP removal from whole blood

100 µL of Pierce™ streptavidin magnetic beads (Thermo Fisher Scientific, USA) was washed three times using 500 µL 1% BSA in PBS before mixing with 3.6 µg of biotinylated anti-CRP mAb and incubated for 1h with mixing at RT. After incubation, beads were washed 5 times using 500 µL 1% BSA in PBST and added to 0.5 mL of whole blood. Capturing was performed at RT for 3 h, before beads were removed using a magnet. CRP removal was evaluated by centrifugation of blood to obtain plasma which was then analyzed by CRP DuoSet ELISA (R&D Systems, USA, no. DY1707) according to manufacturer.

### Multiplexing detection in whole blood

Cross-reactivity of the three sensors was tested by modifying individual electrodes on a single chip with different capturing probes (PCT mAb, APPC and FcMBL protein). PCT (50 ng/ml), CRP (10 µg/ml) or mannan (31.25 ng/mL) were prepared in TBST with 5 mM CaCl2 and 0.1% BSA. 9 chips were prepared in total and 3 were incubated with PCT, 3 with CRP and 3 with mannan. Different biotinylated labels (bio-PCT mAb, bio-CRP mAb and bio-MBL) were mixed and used as a label master mix in all 9 chips to test for cross-reactivity. Multiplexing in whole blood was performed in the same fashion using a mix of biotinylated labels.

## Supporting information

Additional graphs and information to support the manuscript

## Data Availability

Data available on request

## ASSOCIATED CONTENT

### Supporting Information

The following files are available free of charge.

Supplementary material includes comparison of carboxy and amine terminated rGOx on IL-6 sensor sensitivity, antifouling nanocomposite properties in high concentration of Step-Poly HRP, further FE-SEM, AFM and electrochemical characterization of the nanocomposite, sensor validation using SPR, PCT response curve in buffer, demonstration of cross-talk between neighboring electrodes, comparison of infected and non-infected clinical samples using ELISA, PAMPs sensor response in buffer, FcMBL ELISA analysis, mAb based CRP sensor response, PC based CRP sensor in buffer and microfluidic cell set-up for PCT detection (PDF).

## AUTHOR INFORMATION

**Corresponding Author**

*\**.

### Author Contributions

UZ and PJ conceived the study and performed the experiments under the direction of D.M., P.E., and D.E.I.; UZ analyzed the data, which was reviewed by all the authors. All authors contributed to the writing of the manuscript, and all approved the final version.

### Notes

The authors declare no competing financial interests. PJ and DEI are listed as inventors on patents describing this technology.

## ACKNOWLEDGMENTS

The authors would like to thank Drs. M. Super, M. Cartwright, B. Seiler, S. Rifai and N. Shapiro for supplying materials and clinical samples. This work was supported by Defense Advanced Research Projects Agency (DARPA) Contract W911NF-16-C-0050, the Wyss Institute for Biologically Inspired Engineering at Harvard University and the Rosetrees Trust (project M681); the salary of U. Zupan*č*i*č* was provided by University of Bath and Santander. The project was supported by the U.S. Army Research Office, and the content of the information does not necessarily reflect the position or the policy of the Government, and no official endorsement should be inferred.

## Abbreviations

APPC: 4-aminophenyl-phosphorylcholine
AFM: atomic force microscopy
AuNW: gold nanowires
BSA: Bovine serum albumin
CRP: C-reactive protein
CV: cyclic voltammetry
EC: electrochemical
EDC: 1-Ethyl-3-(3-dimethylaminopropyl)carbodiimide
ELISA: enzyme-linked immunosorbent assay
FDA: Food and Drug Administration
FE-SEM: field-emission electron scanning microscope
GA: glutaraldehyde
IL-6: interleukine-6
LOD: limit of detection
MBL: mannose binding lectin
NHS: N-Hydroxysuccinimide
PAMPs: pathogen associated molecular patterns
PBS: phosphate buffer saline
PBST: phosphate buffer saline with 0.5% tween 20
PC: phosphocholine
PCT: procalcitonin
PDMS: polydimethylsiloxane
POC: point-of-care
rGOx: reduced graphene oxide
rGOx-NH2: reduced graphene oxide with amine functionalization
RMS: room mean square
RT: room temperature
SPR: surface plasmon resonance
TBS: Tris-buffered saline
TBST: Tris-buffered saline with 0.5% tween 20
TMB: 3,3’,5,5’-Tetramethylbenzidine
WE: working electrode.

## REFERENCES

1. Kumar, A., Roberts, D., Wood, K. E., Light, B., Parrillo, J. E., Sharma, S., Suppes, R., Feinstein, D., Zanotti, S., Taiberg, L., Gurka, D., Kumar, A., Cheang, M., Duration of hypotension before initiation of effective antimicrobial therapy is the critical determinant of survival in human septic shock. Crit Care Med 2006, 34 (6), 1589–96.

2. Reinhart, K., Bauer, M., Riedemann, N. C., Hartog, C. S., New approaches to sepsis: molecular diagnostics and biomarkers. Clin Microbiol Rev 2012, 25 (4), 609–34.

3. Moschou, D., Tserepi, A., The lab-on-PCB approach: tackling the μTAS commercial upscaling bottleneck. Lab on a Chip 2017, 17 (8), 1388–1405.

4. Hammond, J. L., Formisano, N., Estrela, P., Carrara, S., Tkac, J., Electrochemical biosensors and nanobiosensors. Essays Biochem 2016, 60 (1), 69–80.

5. O’Regan, T. M., Pravda, M., O’Sullivan, C. K., Guilbault, G. G., Development of a disposable immunosensor for the detection of human heart fatty-acid binding protein in human whole blood using screen-printed carbon electrodes. Talanta 2002, 57 (3), 501–10.

6. Bryan, T., Luo, X. L., Bueno, P. R., Davis, J. J., An optimised electrochemical biosensor for the label-free detection of C-reactive protein in blood. Biosens Bioelectron 2013, 39 (1), 94–98.

7. Stern, E., Vacic, A., Rajan, N. K., Criscione, J. M., Park, J., Ilic, B. R., Mooney, D. J., Reed, M. A., Fahmy, T. M., Label-free biomarker detection from whole blood. Nature Nanotechnology 2010, 5 (2), 138–142.

8. Abbott Point of Care Inc. i-STAT® System Test Cartridge Menu. https://www.pointofcare.abbott/int/en/offerings/istat/istat-test-cartridges/menu (accessed Jan 4th,2020).

9. Jolly, P., Formisano, N., Tkáč, J., Kasák, P., Frost, C. G., Estrela, P., Label-free impedimetric aptasensor with antifouling surface chemistry: A prostate specific antigen case study. Sensors and Actuators B: Chemical 2015, 209, 306–312.

10. Ingber, D. E., Henry, O. Y. F., Super, M. Enhanced electrochemical detection using nanoparticles and precipitation. US10753940B2, 2018.

11. Sabate Del Rio, J., Henry, O. Y. F., Jolly, P., Ingber, D. E., An antifouling coating that enables affinity-based electrochemical biosensing in complex biological fluids. Nat Nanotechnol 2019, 14 (12), 1143–1149.

12. Zupancic, U., Jolly, P., Estrela, P., Moschou, D., Ingber, D. E. In Multiplexed Electrochemical Platform for Sepsis Diagnostics, 23rd International Conference on Miniaturized Systems for Chemistry and Life Sciences (Micro), Basel, Switzerland, Basel, Switzerland, 2019.

13. Thangamuthu, M., Hsieh, K. Y., Kumar, P. V., Chen, G.-Y., Graphene- and Graphene Oxide-Based Nanocomposite Platforms for Electrochemical Biosensing Applications. Int J Mol Sci 2019, 20 (12), 2975.

14. Noorbakhsh, A., Alnajar, A. I. K., Antifouling properties of reduced graphene oxide nanosheets for highly sensitive determination of insulin. Microchemical Journal 2016, 129, 310–317.

15. Póvoa, P., Teixeira-Pinto, A. M., Carneiro, A. H., C-reactive protein, an early marker of community-acquired sepsis resolution: a multi-center prospective observational study. Critical Care 2011, 15 (4), R169.

16. Cartwright, M., Rottman, M., Shapiro, N. I., Seiler, B., Lombardo, P., Gamini, N., Tomolonis, J., Watters, A. L., Waterhouse, A., Leslie, D., Bolgen, D., Graveline, A., Kang, J. H., Didar, T., Dimitrakakis, N., Cartwright, D., Super, M., Ingber, D. E., A Broad-Spectrum Infection Diagnostic that Detects Pathogen-Associated Molecular Patterns (PAMPs) in Whole Blood. EBioMedicine 2016, 9, 217–227.

17. Armbruster, D. A., Pry, T., Limit of blank, limit of detection and limit of quantitation. Clin Biochem Rev 2008, 29 Suppl 1, S49–52.

18. Wu, S., Shi, T., Zhang, L., Preparation and properties of amine-functionalized reduced graphene oxide/waterborne polyurethane nanocomposites. High Performance Polymers 2015, 28 (4), 453–465.

19. Potts, J. R., Dreyer, D. R., Bielawski, C. W., Ruoff, R. S., Graphene-based polymer nanocomposites. Polymer 2011, 52 (1), 5–25.

20. Nakatsuka, N., Yang, K. A., Abendroth, J. M., Cheung, K. M., Xu, X., Yang, H., Zhao, C., Zhu, B., Rim, Y. S., Yang, Y., Weiss, P. S., Stojanovic, M. N., Andrews, A. M., Aptamer-field-effect transistors overcome Debye length limitations for small-molecule sensing. Science 2018, 362 (6412), 319–324.

21. Vacic, A., Criscione, J. M., Rajan, N. K., Stern, E., Fahmy, T. M., Reed, M. A., Determination of molecular configuration by debye length modulation. J Am Chem Soc 2011, 133 (35), 13886–9.

22. Arya, S. K., Zhurauski, P., Jolly, P., Batistuti, M. R., Mulato, M., Estrela, P., Capacitive aptasensor based on interdigitated electrode for breast cancer detection in undiluted human serum. Biosensors and Bioelectronics 2018, 102, 106–112.

23. Salimian, R., Kékedy-Nagy, L., Ferapontova, E. E., Specific Picomolar Detection of a Breast Cancer Biomarker HER-2/neu Protein in Serum: Electrocatalytically Amplified Electroanalysis by the Aptamer/PEG-Modified Electrode. ChemElectroChem 2017, 4 (4), 872–879.

24. Fan, S. L., Miller, N. S., Lee, J., Remick, D. G., Diagnosing sepsis - The role of laboratory medicine. Clin Chim Acta 2016, 460, 203–10.

25. Hoeboer, S. H., van der Geest, P. J., Nieboer, D., Groeneveld, A. B. J., The diagnostic accuracy of procalcitonin for bacteraemia: a systematic review and meta-analysis. Clinical Microbiology and Infection 2015, 21 (5), 474–481.

26. Kondo, Y., Umemura, Y., Hayashida, K., Hara, Y., Aihara, M., Yamakawa, K., Diagnostic value of procalcitonin and presepsin for sepsis in critically ill adult patients: a systematic review and meta-analysis. J Intensive Care 2019, 7, 22.

27. Kozel, T. R., Burnham-Marusich, A. R., Point-of-Care Testing for Infectious Diseases: Past, Present, and Future. Journal of Clinical Microbiology 2017, 55 (8), 2313.

28. von Heimburg, D., Stieghorst, W., Khorram-Sefat, R., Pallua, N., Procalcitonin—a sepsis parameter in severe burn injuries. Burns 1998, 24 (8), 745–750.

29. Uusitalo-Seppälä, R., Koskinen, P., Leino, A., Peuravuori, H., Vahlberg, T., Rintala, E. M., Early detection of severe sepsis in the emergency room: diagnostic value of plasma Creactive protein, procalcitonin, and interleukin-6. Scandinavian journal of infectious diseases 2011, 43 (11-12), 883-890.

30. O’Grady, N. P., Barie, P. S., Bartlett, J. G., Bleck, T., Carroll, K., Kalil, A. C., Linden, P., Maki, D. G., Nierman, D., Pasculle, W., Masur, H., Guidelines for evaluation of new fever in critically ill adult patients: 2008 update from the American College of Critical Care Medicine and the Infectious Diseases Society of America. Crit Care Med 2008, 36 (4), 1330–49.

31. Ren, H., Li, Y., Han, C., Hu, H., Serum procalcitonin as a diagnostic biomarker for sepsis in burned patients: a meta-analysis. Burns 2015, 41 (3), 502–9.

32. Kuse, E. R., Langefeld, I., Jaeger, K., Kulpmann, W. R., Procalcitonin in fever of unknown origin after liver transplantation: a variable to differentiate acute rejection from infection. Crit Care Med 2000, 28 (2), 555–9.

33. Liu, D., Su, L., Han, G., Yan, P., Xie, L., Prognostic Value of Procalcitonin in Adult Patients with Sepsis: A Systematic Review and Meta-Analysis. PLoS One 2015, 10 (6), e0129450.

34. Liu, Y., Hou, J.-h., Li, Q., Chen, K.-j., Wang, S.-N., Wang, J.-m., Biomarkers for diagnosis of sepsis in patients with systemic inflammatory response syndrome: a systematic review and meta-analysis. SpringerPlus 2016, 5 (1), 2091.

35. Wirz, Y., Meier, M. A., Bouadma, L., Luyt, C. E., Wolff, M., Chastre, J., Tubach, F., Schroeder, S., Nobre, V., Annane, D., Reinhart, K., Damas, P., Nijsten, M., Shajiei, A., deLange, D. W., Deliberato, R. O., Oliveira, C. F., Shehabi, Y., van Oers, J. A. H., Beishuizen, A., et al., Effect of procalcitonin-guided antibiotic treatment on clinical outcomes in intensive care unit patients with infection and sepsis patients: a patient-level meta-analysis of randomized trials. Crit Care 2018, 22 (1), 191.

36. Dolin, H. H., Papadimos, T. J., Stepkowski, S., Chen, X., Pan, Z. K., A Novel Combination of Biomarkers to Herald the Onset of Sepsis Prior to the Manifestation of Symptoms. Shock 2018, 49 (4), 364–370.

37. Shapiro, N. I., Trzeciak, S., Hollander, J. E., Birkhahn, R., Otero, R., Osborn, T. M., Moretti, E., Nguyen, H. B., Gunnerson, K. J., Milzman, D., Gaieski, D. F., Goyal, M., Cairns, C. B., Ngo, L., Rivers, E. P., A prospective, multicenter derivation of a biomarker panel to assess risk of organ dysfunction, shock, and death in emergency department patients with suspected sepsis. Society of Critical Care Medicine 2009, 37 (1).

38. Reddy, B., Hassan, U., Seymour, C., Angus, D. C., Isbell, T. S., White, K., Weir, W., Yeh, L., Vincent, A., Bashir, R., Point-of-care sensors for the management of sepsis. Nature Biomedical Engineering 2018, 2 (9), 640–648.

39. Kang, J. H., Super, M., Yung, C. W., Cooper, R. M., Domansky, K., Graveline, A. R., Mammoto, T., Berthet, J. B., Tobin, H., Cartwright, M. J., Watters, A. L., Rottman, M., Waterhouse, A., Mammoto, A., Gamini, N., Rodas, M. J., Kole, A., Jiang, A., Valentin, T. M., Diaz, A., Takahashi, K., Ingber, D. E., An extracorporeal blood-cleansing device for sepsis therapy. Nat Med 2014, 20 (10), 1211–6.

40. Seiler, B. T., Cartwright, M., Dinis, A. L. M., Duffy, S., Lombardo, P., Cartwright, D., Super, E. H., Lanzaro, J., Dugas, K., Super, M., Ingber, D. E., Broad-spectrum capture of clinical pathogens using engineered Fc-mannose-binding lectin enhanced by antibiotic treatment. F1000Res 2019, 8, 108.

41. Ugarte, H., Silva, E., Mercan, D., De Mendonca, A., Vincent, J. L., Procalcitonin used as a marker of infection in the intensive care unit. Crit Care Med 1999, 27 (3), 498–504.

42. Celik, I. H., Demirel, F. G., Uras, N., Oguz, S. S., Erdeve, O., Biyikli, Z., Dilmen, U., What are the cut-off levels for IL-6 and CRP in neonatal sepsis? J Clin Lab Anal 2010, 24 (6), 407–12.

43. Tanaka, T., Robey, F. A., A new sensitive assay for the calcium-dependent binding of Creactive protein to phosphorylcholine. Journal of Immunological Methods 1983, 65 (3), 333–341.

44. Thompson, D., Pepys, M. B., Wood, S. P., The physiological structure of human Creactive protein and its complex with phosphocholine. Structure 1999, 7 (2), 169–177.

45. Christopeit, T., Gossas, T., Danielson, U. H., Characterization of Ca2+ and phosphocholine interactions with C-reactive protein using a surface plasmon resonance biosensor. Analytical Biochemistry 2009, 391 (1), 39–44.

46. Buchegger, P., Preininger, C., Four Assay Designs and On-Chip Calibration: Gadgets for a Sepsis Protein Array. Analytical Chemistry 2014, 86 (6), 3174–3180.

