## Additional graphs and information to support the manuscript for "Multiplexed Detection of Sepsis Markers in Whole Blood using Nanocomposite Coated Electrochemical Sensors"

SUPPORTING INFORMATION FOR PUBLICATION

*Performance of the IL-6 sensor*

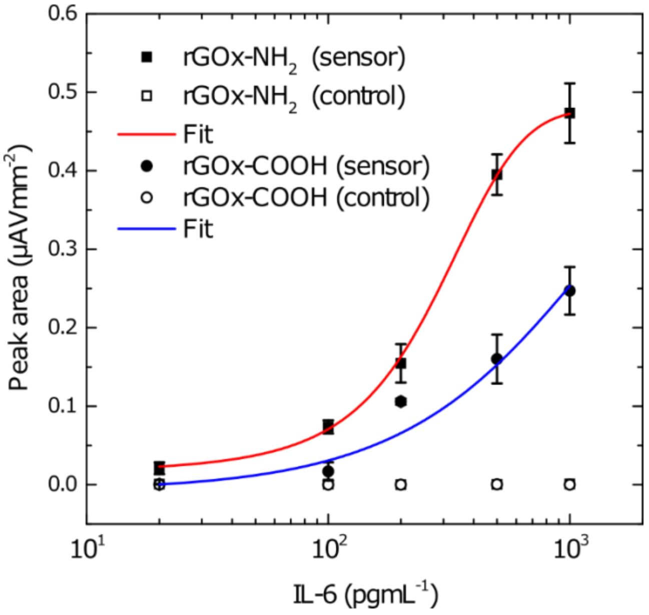


**Figure S1.** Electrochemical IL-6 sensors in undiluted serum based on nanocomposite with amine terminated rGOx and carboxyl terminated rGOx. Full symbols represent the average (n = 3) and error bars the standard deviation. Hill fit was performed to fit the data. Empty symbols represent the response obtained using non-specific, IL-8 capturing mAb. The IL-6 capturing antibody was conjugated to the nanocomposite using EDC/NHS chemistry. After conjugation, the surface was washed with PSB for 5 min while shaking and quenched with 1M ethanolamine in PBS for 1h. Then blocking was performed with 1%BSA solution in PBST for minimum of 1h. IL-6 was thawed on ice and diluted in human serum before 10 µL of the sample was deposited on each chip and incubated for 1h, followed by 30min incubation of detection antibody at 10 µgmL$^{-1}$, 5 min incubation of Streptavidin-PolyHRP conjugate (1 μgmL$^{-1}$) and 1 min incubation with precipitating TMB.

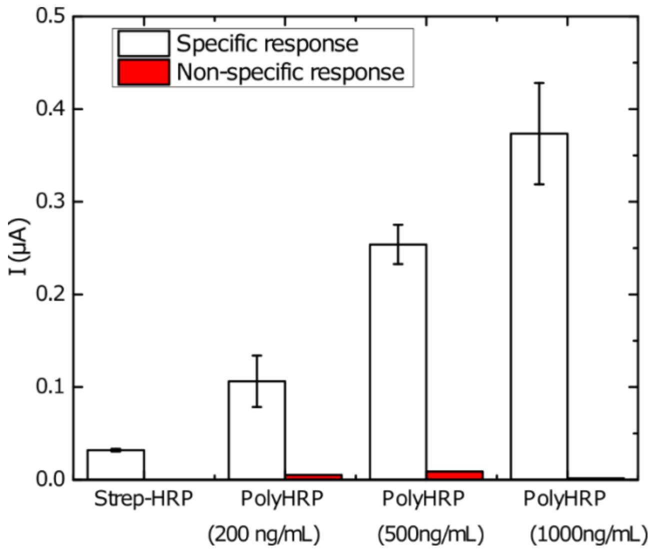


**Figure S2.** Increasing concentrations of streptavidin-PolyHRP for effective signal enhancement. Empty bars represent the average (n = 3) and error bars the standard deviation. Specific response refers to electrodes with IL-6 capturing mAb and non-sepcific response to electrodes with IL-8 capturing mAb. IL-6 sensors were constructed as described above and 200 pgmL$^{-1}$ target concentration was used in all experiments in **Figure S2**. Streptavidin-HRP conjugate was used (recommended concentration according to manufacturer's protocol) along with Streptavidin-PolyHRP conjugate with increasing concentrations. As demonstrated, the signal increased with increasing enzyme concentration and a minimal non-specific response can be observed due to antifouling properties of the nanocomposite.

*Nanocomposite characterization*

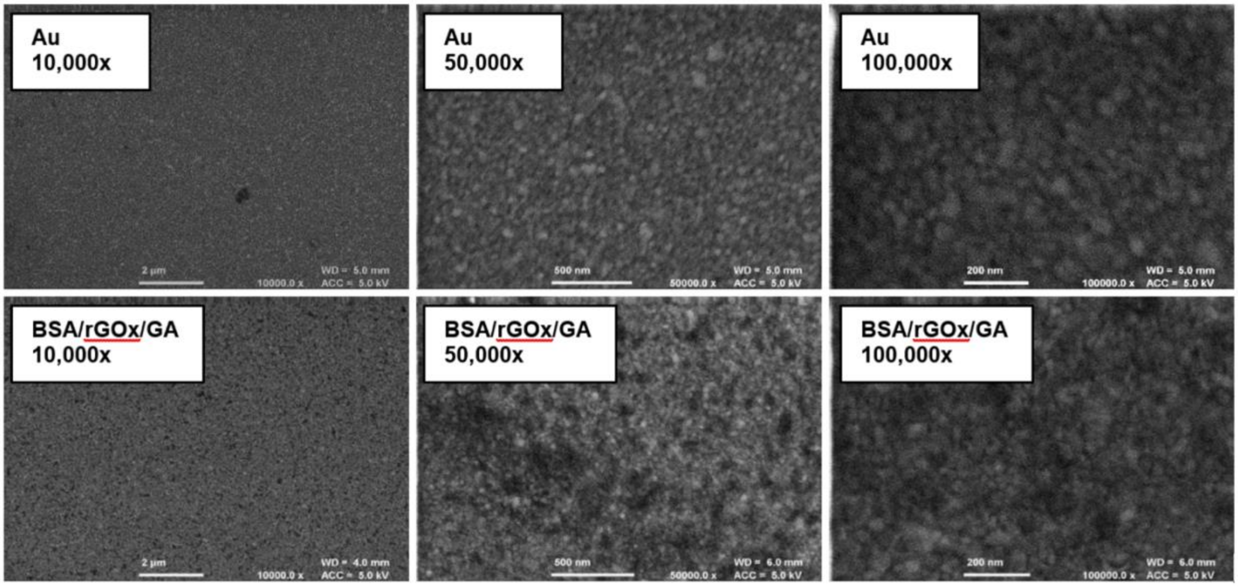


**Figure S3.** FE-SEM imaging of BSA/rGOx/GA nanocomposite. Bare gold in the top row and BSA/rGOx/GA nanocomposite in the bottom row at magnifications from 10,000x to 100,000x.

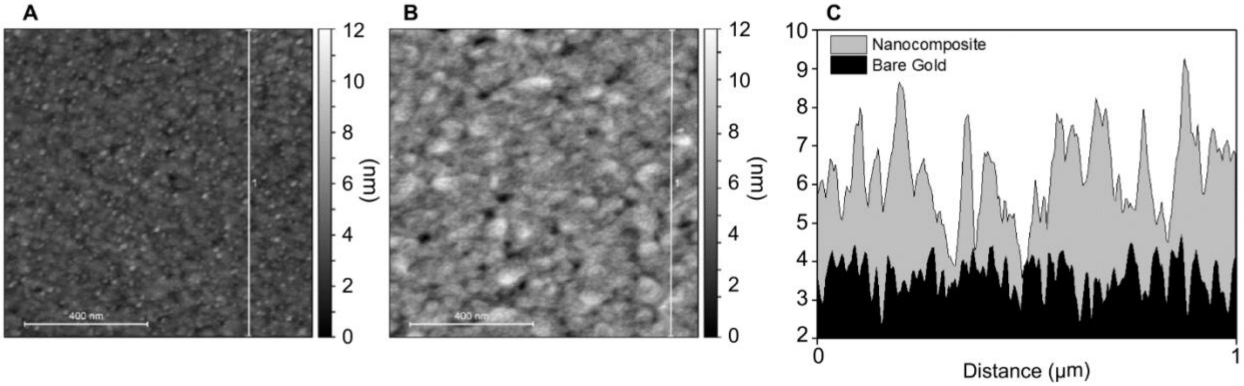


**Figure S4.** AFM analysis of bare gold and BSA/rGOx/GA nanocomposite. (A) Bare gold scan from which the profile was extracted. (B) BSA/rGOx/GA nanocomposite image from which the profile was extracted. (C) The extracted profiles of both samples.

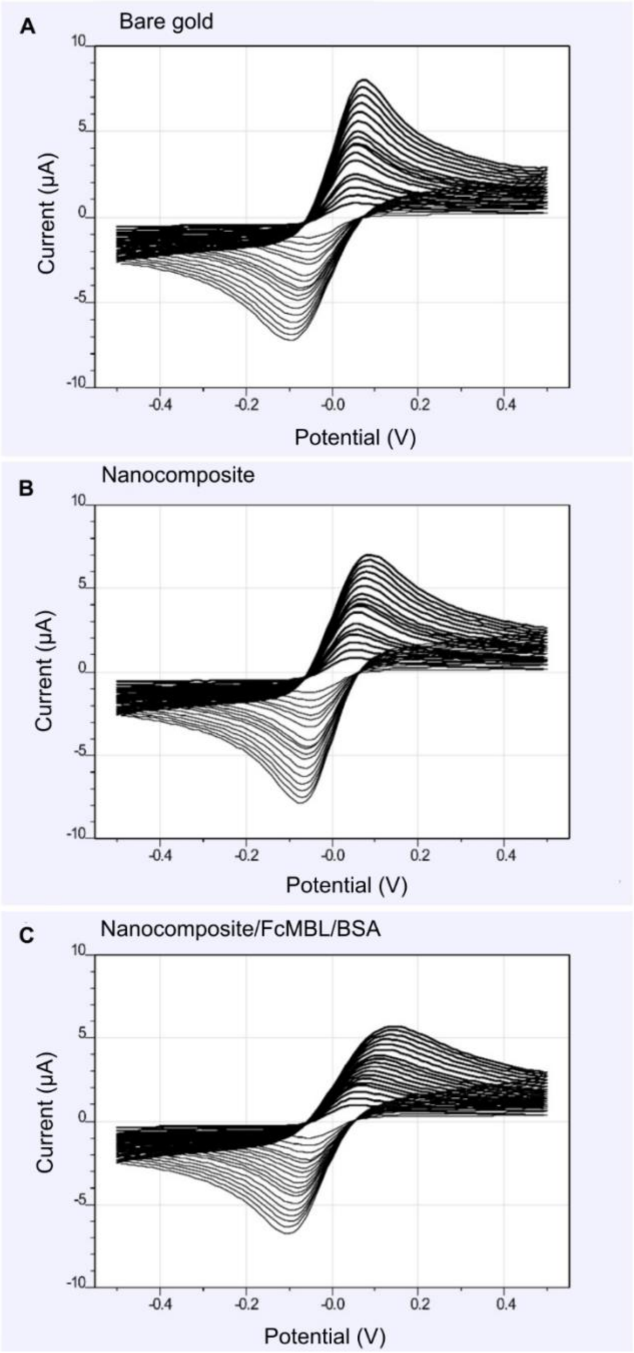


**Figure S5.** Oxidation and reduction current obtained in 5 mM ferri-/ferrocyanide using various scan rates between 0.01 V/s to 1 V/s on bare gold (A) BSA/rGOx/GA nanocomposite (B) and FcMBL functionalized nanocomposite after blocking with BSA (C).

*Procalcitonin sensor characterization and validation using surface plasmon resonance*

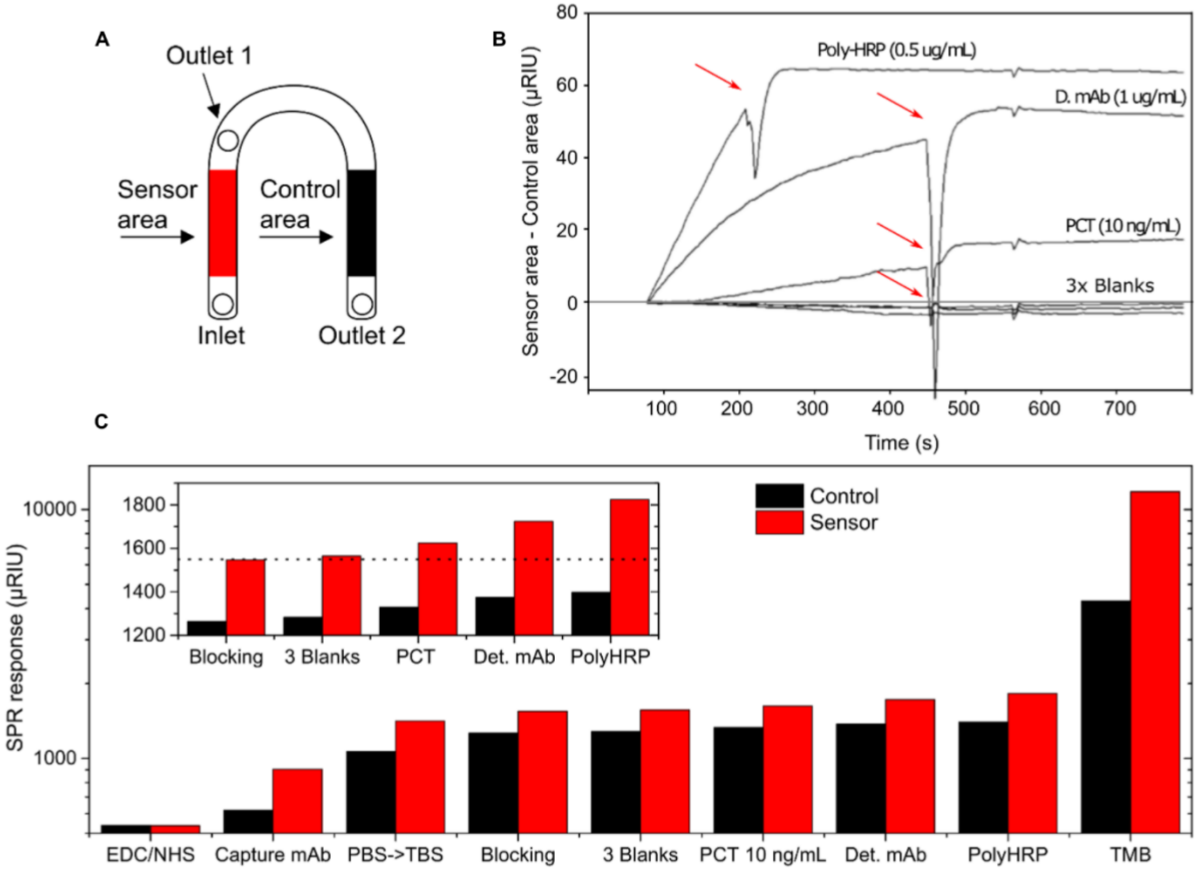


**Figure S6**. Validation of the PCT assay using SPR. Schematic representation of the SPR flow cell used in the study (A). Raw data obtained from the injection of blanks, PCT sample, detection mAb and Strep-pHRP. Red arrows indicate the transition between association and dissociation step (B). Combined data from sensor construction through pTMB precipitation. Inset presents closeup from blocking to Strep-pHRP steps (C).

Gold SPR chips were dipped in hot piranha solution for 5 min and cleaned with oxygen plasma system at 100 W, 0.5 mbar for 5 min before the nanocomposite was formed on the gold as described. The following day, the chip was washed for 30 min and inserted in a dual channel SR700DCSPR system (Reichert Technologies, Germany). The SPR flow cell allowed us to specifically functionalize one side of the channel, leaving the second side as a control. Nanocomposite was then activated by flowing EDC/NHS solution from inlet to outlet 2, see **Figure S6a**. Capturing PCT mAb was then flown from inlet to outlet 1, leaving the control area non-functionalized. Both channels were then quenched and blocked. **Figure S6b** represents the difference between the sensor area and the control area after the injection of PCT, det. mAb and Strep-pHRP. The increase in the SPR response indicates specific binding, which is increasing in the association phase and remains constant in the dissociation phase. **Figure S6c** reveals characterization of every step in EC sensor construction. After surface activation, capturing mAb is specifically conjugated to the sensing area. Quenching and blocking steps do not significantly change the surface which remains stable after the injection of three blanks. Injection of PCT, detection mAb and Strep-pHRP can be seen as a signal increase. After the injection of pTMB, very big increase can be seen due to precipitation of the pTMB to the surface, significantly changing surface properties. Due to the downstream positioning of the control channel some signal increase can also be observed in the control area.

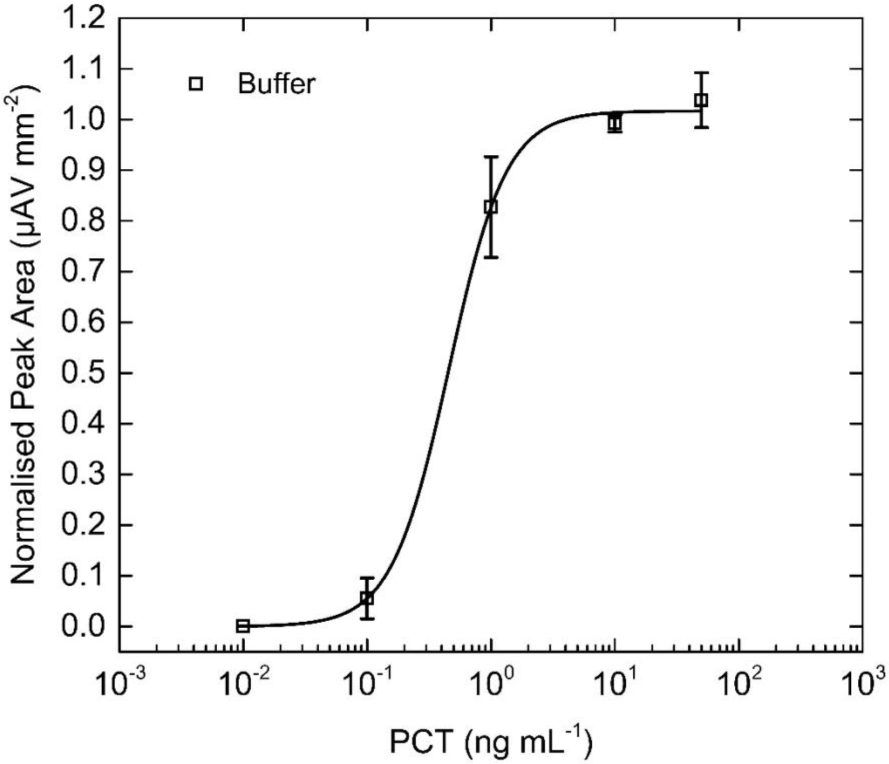


**Figure S7.** Electrochemical PCT sensor response in in buffer. Squares represent the average (n = 3) and error bars the standard deviation. Hill fit was performed to fit the data.

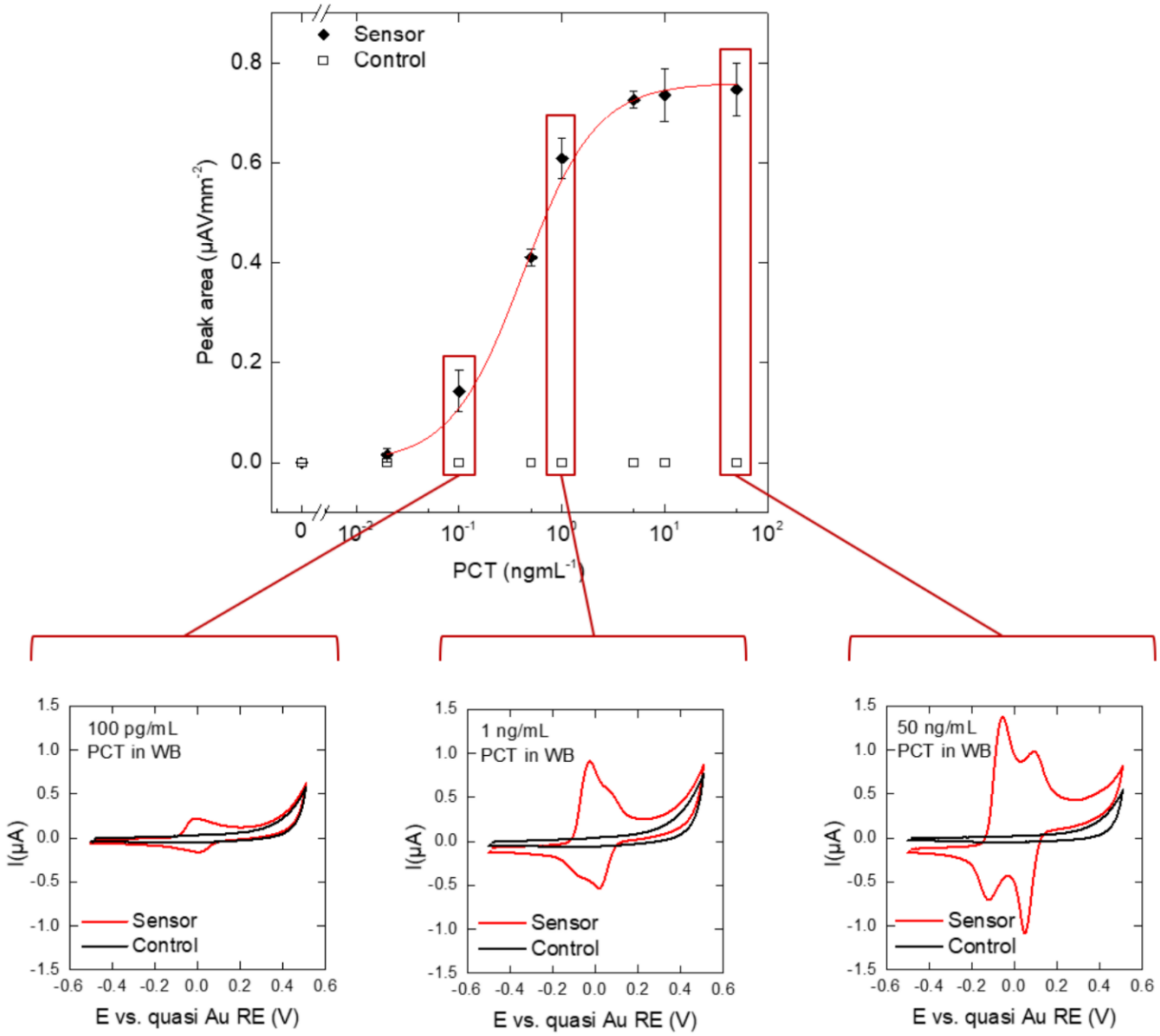


**Figure S8.** PCT sensor in 50% diluted whole blood. Plot on the top represents calibration curve obtained by increasing the PCT concentrations in blood. Full symbols represent the average (n = 3) and error bars the standard deviation. Hill fit was performed to fit the data. Empty circles represent the response obtained using the control electrode with no capturing mAb. Plots in the bottom row represent the response from two neighboring electrodes, first bearing anti-PCT mAb (red) and second with no capturing probe (black) at 0.1 ngmL$^{-1}$, 1 ngmL$^{-1}$ and 50 ngmL$^{-1}$ concentration of PCT in the sample.

*Clinical sample analysis using plate ELISA*

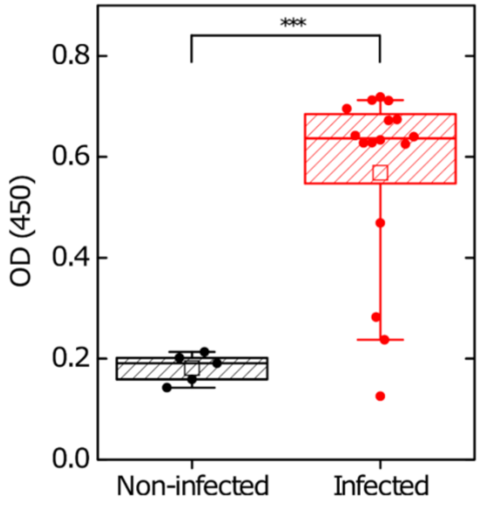


**Figure S9.** Box plot comparing PCT levels in infected and non-infected patient groups obtained with plate ELISA. Full circles represent the mean of measured PCT values, lines in a box represent median, $25^{th}$ and $75^{th}$ percentile, empty square is mean value, whiskers represent $10^{th}$ and $90^{th}$ percentile and *** represents $p < 0.001$.

*Pathogen associated molecular patterns sensor characterization*

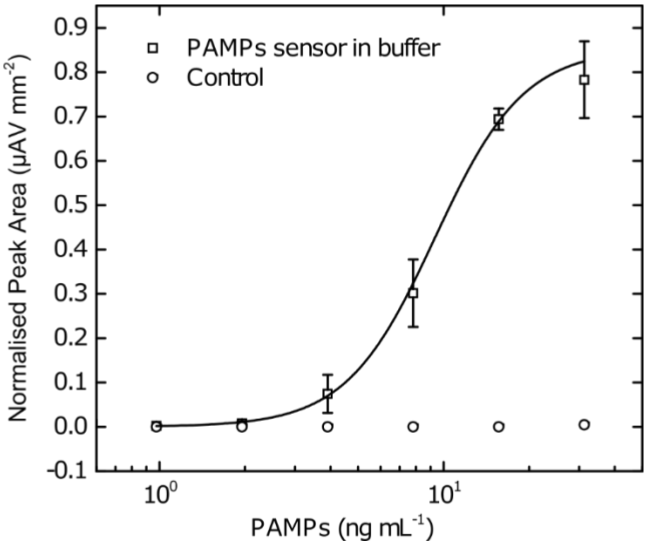


**Figure S10.** Electrochemical PAMPs sensor in buffer. Mannan was added to in increasing concentrations, to obtain the response curve. Squares represent the average (n = 3) and error bars the standard deviation while empty circles represent the response obtained using the control electrode with no capturing probe. Hill fit was performed to fit the data

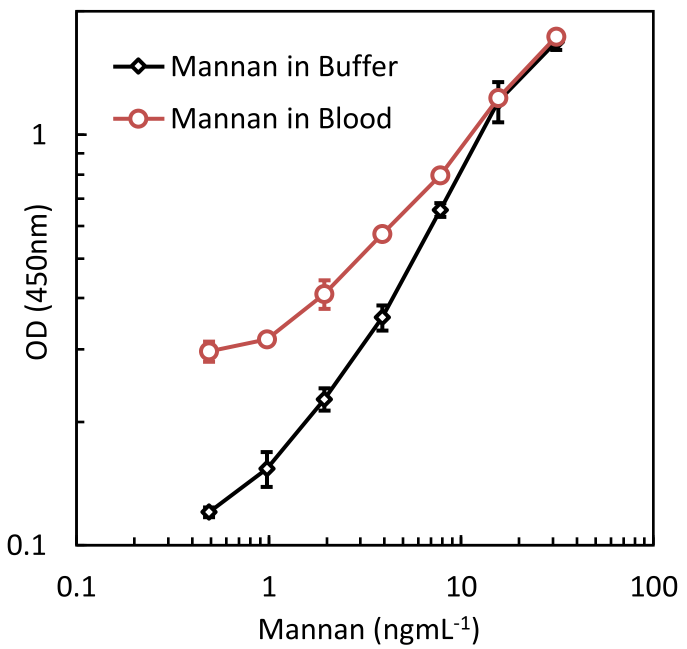


**Figure S11.** FcMBL based ELISA on plate with increasing mannan concentrations. Comparison of response in buffer and 50% blood. Symbols represent the average (n = 3) and error bars the standard deviation. As observed in the EC sensor, higher background can be observed in the assay with blood demonstrating some PAMPs are present in the biological sample.

*Depletion of CRP in blood*

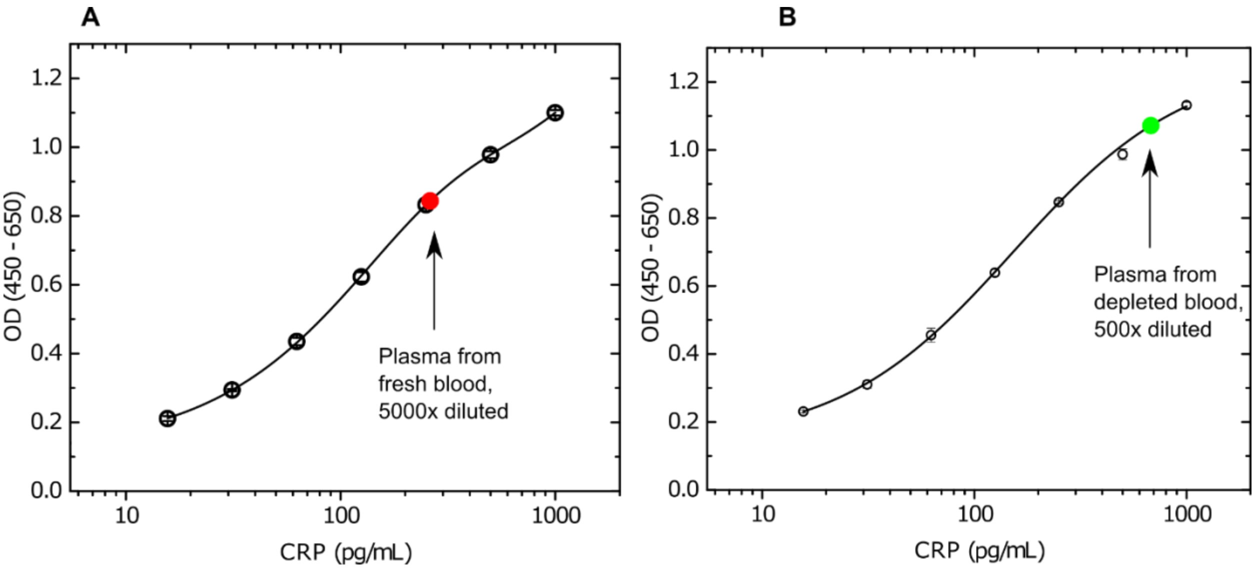


**Figure S12**. CRP ELISA on plate demonstrating depletion of CRP level in plasma from whole blood after CRP capturing using magnetic beads. The plasma was diluted in PBS and analyzed using CRP DuoSet ELISA according to manufacturer.

*C-reactive protein sensor*

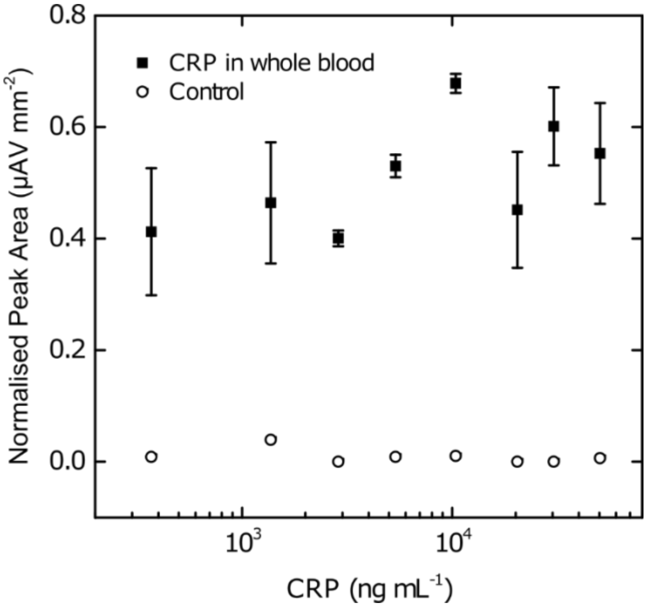


**Figure S13.** Antibody based electrochemical CRP sensor in 50% whole blood with depleted CPR level. Squares represent the average (n = 3) and error bars the standard deviation while empty circles represent the response obtained using the control electrode with no capturing probe.

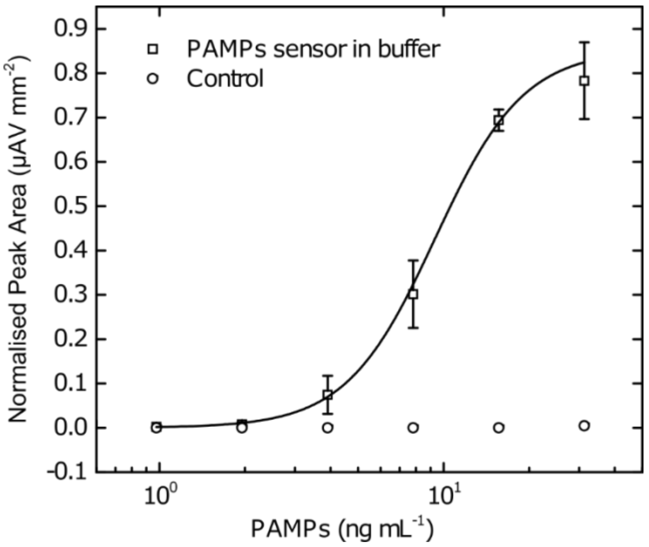

**Figure S14**. Electrochemical CRP sensor based on phosphocholine in buffer. CRP is added to the blocking buffer to obtain the calibration curve. Squares represent the average (n = 3) and error bars the standard deviation. Datapoint at 100 ngmL$^{-1}$ CRP includes only one measurement. Hill fit was used to fit the data. Hill fit was performed to fit the data.

*Microfluidic integration of the procalcitonin sensor*

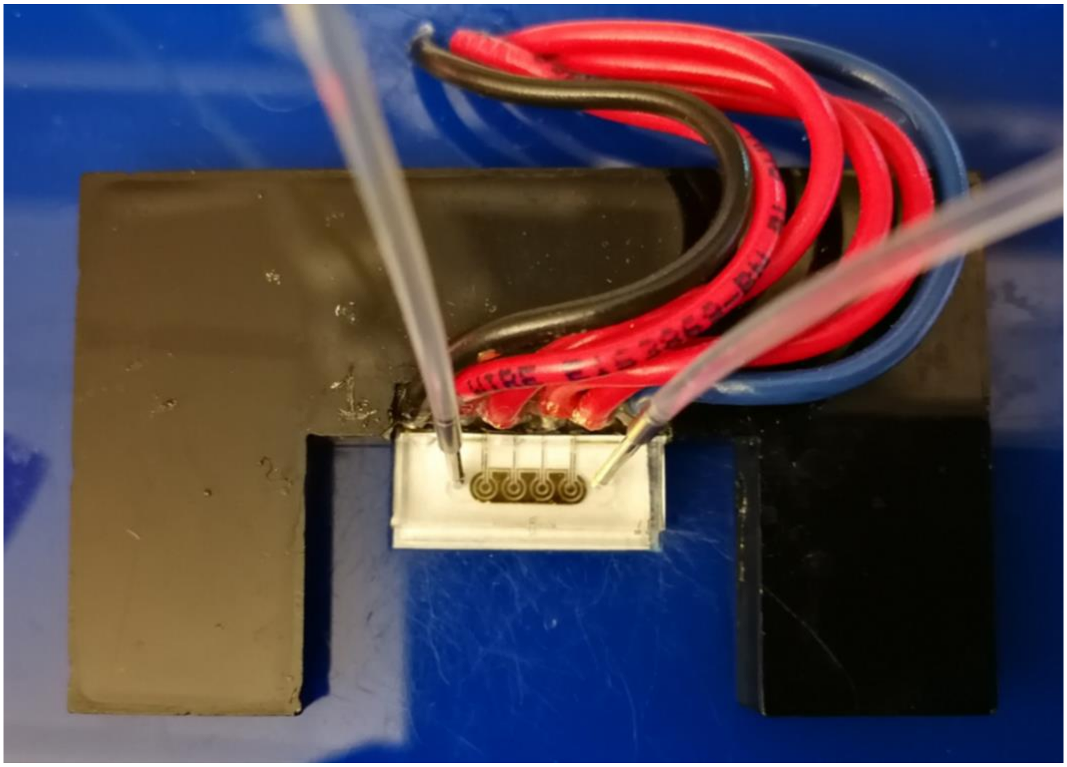

**Figure S15.** Picture of a microfluidic cell attached on the nanocomposite covered chip. Double sided adhesive was used to outline the channel and a PDMS block with an inlet and outlet holes is adhered on the top. Stainless steel connectors are inserted in the PDMS connecting the tubing to the microfluidic chamber.
